# Predicting the Trajectory of Any COVID19 Epidemic From the Best Straight Line

**DOI:** 10.1101/2020.06.26.20140814

**Authors:** Michael Levitt, Andrea Scaiewicz, Francesco Zonta

## Abstract

A pipeline involving data acquisition, curation, carefully chosen graphs and mathematical models, allows analysis of COVID-19 outbreaks at 3,546 locations world-wide (all countries plus smaller administrative divisions with data available). Comparison of locations with over 50 deaths shows all outbreaks have a common feature: *H(t)* defined as log_e_*(X(t)/X(t-1))* decreases linearly on a log scale, where *X(t)* is the total number of Cases or Deaths on day, *t* (we use ln for log_e_). The downward slopes vary by about a factor of three with time constants (1/slope) of between 1 and 3 weeks; this suggests it may be possible to predict when an outbreak will end. Is it possible to go beyond this and perform early prediction of the outcome in terms of the eventual plateau number of total confirmed cases or deaths?

We test this hypothesis by showing that the trajectory of cases or deaths in any outbreak can be converted into a straight line. Specifically *Y*(*t*) ≡ −ln(ln(*N* / *X* (*t*)), is a straight line for the correct plateau value *N*, which is determined by a new method, Best-Line Fitting (BLF). BLF involves a straight-line facilitation extrapolation needed for prediction; it is blindingly fast and amenable to optimization. We find that in some locations that entire trajectory can be predicted early, whereas others take longer to follow this simple functional form. Fortunately, BLF distinguishes predictions that are likely to be correct in that they show a stable plateau of total cases or death (*N* value). We apply BLF to locations that seem close to a stable predicted *N* value and then forecast the outcome at some locations that are still growing wildly. Our accompanying web-site will be updated frequently and provide all graphs and data described here.

## INTRODUCTION

In December 2019 a coronavirus, known as SARS-CoV-2, was discovered in Wuhan China (Wang, 2020). The virus, perhaps from horseshoe bats (Zhou, 2020), spread between humans during January 2020, leading to the COVID-19 pandemic. Early prediction of the number of cases and deaths in an epidemic or pandemic is of vital importance as it helps policy makers make informed decisions on the best allocation of resources and containment of the pathogen. For this reason, many different groups have attempted to make reliable predictions of Sars-Cov-2 diffusion (Levitt 2020a, Wang 2020, Dimeglio 2020, Wu 2020, Pinotti 2020). These forecasts are based on a variety of mathematical and statistical models, which use different types of data (COVID-19 data, mobility data, demographic data) and take into account the impact of interventions, such as social distancing, proper hand hygiene and the use of masks. Such variables differ from country to country, and moreover, the criteria to detect COVID19 cases and consider COVID19 as the cause of deaths also vary sometimes even for states/provinces in the same country. These factors combine to complicate finding a universal method to fit and predict COVID-19 trajectories.

We began working on COVID-19 in the last week of January 2020 using data released by Sudalai Rajkumar (Rajkumar), Johns Hopkins Coronavirus Resource Center (JHCS) and Chinese internet (JOBTUBE). On January 28^th^, there were numbers of cases and deaths for 6 days starting on January 22^nd^. The daily death rate of COVID-19 (ratio of total deaths to total cases on a given day) was ten times higher inside Hubei, the province surrounding Wuhan, than everywhere else in China (non-Hubei). Concerned and encouraged by this data, we started an Excel spreadsheet to follow the daily progression of COVID-19. Each day, we made graphs of four simple measures. Three were obvious: the total number of cases; the total number of deaths, and their ratio, the death rate. The fourth was trivial but less obvious: the ratio of the total cases (or deaths) denoted as *X(t)* for today divided by that of yesterday. This ‘fractional change function’ *f* (*t*) measures exponential growth of *X(t)* with *f* (*t*) = *X* (*t*) / *X* (*t* − 1).

If the total today is always 10% more than yesterday the value today will be 1.1 times the value yesterday with *f(t)=1*.*1*. In fact, on January 29^th^, the number of deaths today divided by that of yesterday was 1.3. Were such exponential growth of 30% a day to continue, everyone on earth would die within 90 days. Analyzing the data more completely over the next few days, we noticed on February 2^nd^ that the fractional change for deaths in Hubei showed a steady decrease from 30% on January 29^th^ to 18% four days later. If this linear decrease of fractional change in deaths continued then deaths in Hubei would stop on day 67, when the fractional change became equal 1 can *X(t)* was the same as *X(t-1)*. We reported this finding widely (Levitt, 2020b), although in retrospect, it was naïve to expect the linear decrease to continue.

Nevertheless, our early interest in the fractional change function remained for two reasons. Firstly, because of the mathematical simplicity of *X* (*t*) / *X* (*t* − 1) as compared to more accepted measures like *R*_*t*_(Wallinga 2007; Ferguson 2013). Secondly, because, by analyzing the data of a small number of early epidemics (before mid-March 2020), we realized that the factional change function appears to have the same shape for multiple locations: it converges to 1 as fast as a decaying exponential (Levitt, 2020c). Furthermore, because the fractional change function is a ratio, it is not affected by different systematic counts of cases/deaths due to different criteria: two countries that apply different criteria for deciding when a person is infected but have the same day to day growth will have the same fractional change function, provided that the counting method is kept consistent within the country.

Elaborating further from this initial intuition we found a minimal mathematical model that allows us to consistently describe the spreading of the virus in different countries. We also were able to reduce the very complicated task of fitting inconsistent data sets to the fitting of a straight line for which extrapolations and quality controls are trivial. This allowed us to completely automate data fitting, extrapolation and assessment of the quality of fitting, all done simultaneously and at blinding speed (less than an hour of CPU for all the outbreaks in the world).

## METHODS

### Data Processing

Data is synced daily to two different sources for world data, US and Italy data. World data and US data including county and states levels is taken from (JHU), available from (Starschema). Italy data at provinces level is taken from (Ita-regioni). (We thank Levitt-group members Dr. João Rodrigues and Dr. Frederic Poitevin for integrating these data sources into a master file).

For some location the data contains inconsistencies, which we call ‘data glitches’ and these are corrected as we did in our earliest analysis of the epidemic in Hubei, China (Levitt, 2020d). We were well-aware that any alteration of the raw data must be justified and carefully recorded as we do here. Such correction turns out to be important as the curve-fitting of the raw data is insensitive to random counting errors, allowing us to use the raw data without any smoothing, but is sensitive to systematic errors like these. There were three type of ‘data glitches’: ‘mis-glitches’, ‘rise-glitches’ and ‘drop-glitches’. (1) *mis-glitches* occur when the data on a given day is not updated. Specifically, whenever two consecutive *X(t)* values (at times *t-1* & t) are identical, we alter the value at *t* to be the average of the values at times *t-1* & *t+1*. (2) *rise-glitches* occur when new cases or deaths not previously reported are discovered and released on a particular day. This first occurred in China Hubei on February 13^th^, when 13,000 cases detected clinically were added to the total. These cases did not occur on the day reported but rather over the preceding days, so we corrected for by rescaling the number of confirmed cases on days prior to 13^th^ February by a constant factor greater than 1 (Levitt 2020d). The same correction was applied on a small number of instances when additional deaths or confirmed cases were reported on a specific day as having been unreported on previous days. Again, we added the deaths or confirmed cases to the previous days a fixed fractional increment (the complete list of with both types of correction is provided in the Supplementary Material). (3) *drop-glitches* occur when the total numbers at a given location are decreased on a particular day. This can never happen normally as totals always increase and is due to the realization that numbers reported previously include misidentified cases or deaths. This glitch is less common than the other two. It is corrected in the same way as the *rise-glitch* except that the factor multiplying total values on all previous days is less than 1.

### Mathematical background

We consider *X* (*t*), the discrete temporal series of cases (or deaths) in a given country, region or province. In the most general scenario, we assume that *X* (*t*) obeys the following ordinary differential equation (ODE):

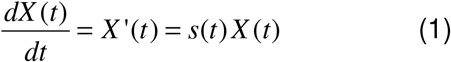

In the discretized form the first derivative of *X(t)* is *X* ‘(*t*) = *X* (*t* − 1) − *X* (*t*), which is the number of new cases on day *t*. Equation (1) simply states that the number of new cases on a certain day is proportional to the number of cases on the previous day.

The coefficient of proportionality *s(t)* is not constant. It changes with time so as to take into account the dynamics of virus spreading, which may be affected by social distancing or the structure of social network interactions.

We are interested in a solution of Equation 1 that reaches a plateau value of *N* for a large *t* (often called a growth function). A general form for many different kinds of growth functions can be written as follows (Koya 2013)

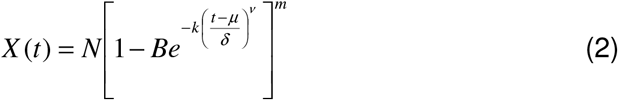

Equation 2 describes a rich family of curves which comprises Richards functions (Richards 1959), generalized logistic functions, Weibull functions (Frechet 1927) etc. While the overall shape of these curves depends on the various parameters, the asymptotic behavior has the same analytical form for all the curves in the family. It is this behavior that allows us to introduce an important simplification that reduces the fitting of Equation 2 to fitting a straight line. It is easy to show that the following relationship holds in the limit of large *t*:

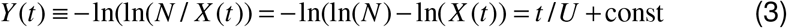

Equation (3) is true asymptotically for every function in the Koya Goshu family, and exactly true for the Gompertz function, *G(t)*, (Gompertz 1825). This function has been also used by other groups to fit data of COVID-19 trajectories (Castorina 2020, Catala 2020) and is shown in **Fig 1** and **Fig. S1**):

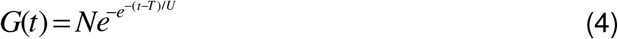

**Figure 1.**
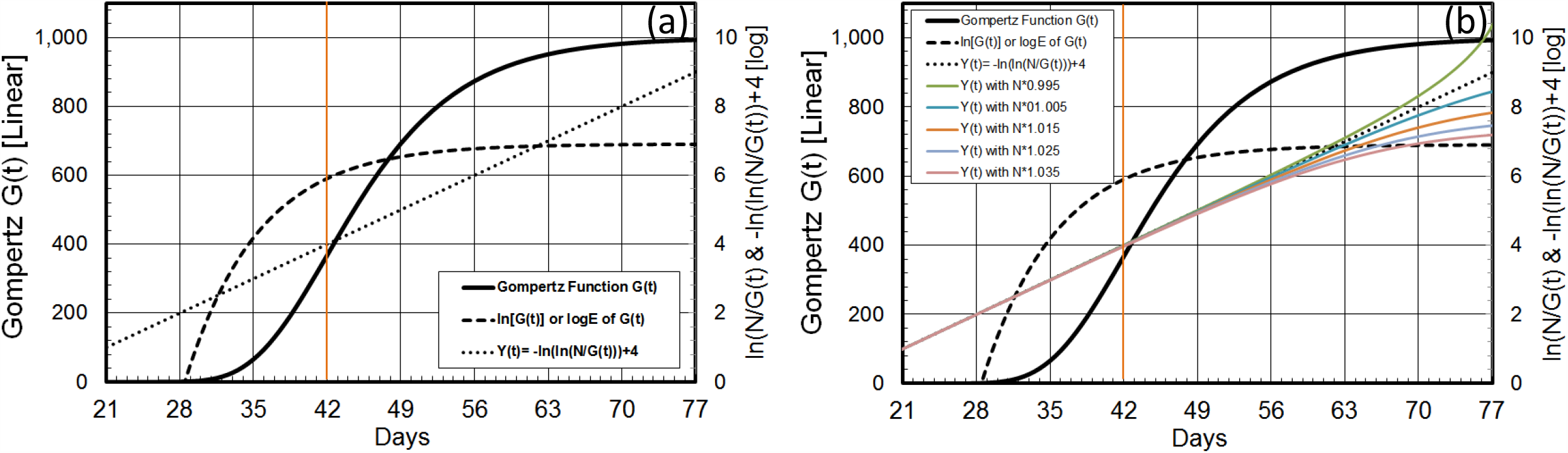
Showing the Gompertz Function *Y(t)* Straightened to Line Y(y) to Predict Plateau *N*. (a) Basic properties of the Gompertz functions and its logarithms. The Gompertz function is an exponential of an exponential written as 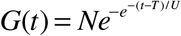 or *G*(*t*) = *N* exp(− exp(− (*t* − *T*) / *U*)), and defined by three parameters *N, T* & *U*, each with clear physical meaning. Parameter *N* is the asymptotic number, the maximum plateau value that *G(t)* reaches after a long time, *t*. Parameter *T*, is the point of inflection, which is the time in days at which the second-derivative of *G(t)* is zero and its first derivative is a maximum. It is a natural mid-point of the function where the value *of G(T)=N/e=0*.*37N*. The Parameter *U*, is the most important as it changes the shape of the curve; it is a time-constant measured in days. Given the double exponential nature of *G(t)*, one might expect to use a double logarithm to simplify it. The function *G(t)* itself has the expected S-shape of saturating growth function. Taking the logarithm once gives ln(*G*(*t*)) = ln(*N*)−exp(− (*t* −*T*)/*U*), where ln is the natural logarithm or *log*_*e*_; it is shown in dashed line increasing very rapidly at first but curving steadily to become horizontal at saturation. Rearranging as ln(*N*)−ln(*G*(*t*)) = exp(− (*t* −*T*)/*U*)and taking the logarithm a second time gives *Y* (*t*) = −ln[ln(*N*)− ln(*G*(*t*))] = −ln[ln(*N* / *G*(*t*))]= (*t* −*T*) /*U*. This function is shown in the dotted line to be a simple straight line. This is hugely significant as extrapolation of a straight-line is trivial: just keep going straight. As we show in the text, the function *Y(t)* is always a straight line for the Gompertz function. More generally, *Y(t)*, tends to a straight line for a very general class of saturating functions (b) Illustrating how the linearity of the *Y(t)=-ln(ln(N/G(t))* depends on the value of *N*. The linearity shown in (a) has an apparent weakness, namely the line is only straight when the value of *N* is the correct saturation value and this value will be unknown until the epidemic is over. This weakness is in fact a strength. One can try different values of *N* and find the one that gives a straight line. In fact, “straighten the line” is much more relevant than the saying “flatten the curve” popularly applied to COVID19.

We also consider another function, which is the logarithm of the fractional change function *f* (*t*) defined above:

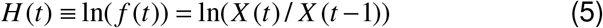

For a Gompertz distribution *H(t)* is a decaying exponential function with the same time constant *U*, associated with *Y(t):*

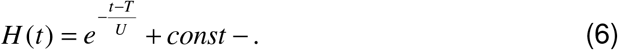

A similar relationship is valid asymptotically for other growing functions with the same time constant.so analysis of the behavior of *H(t)*, provides a second method to derive the time constant *U*.

### Data fitting and validation

The simple linear relationship in Equation 3 provides a remarkable tool allowing us to fit the trajectory of virus spreading and predict the end points (*N*) in different locations. Given a single data series *X(t)*, the best estimate for *ln(N)* is determined as the value that maximizes the correlation coefficient of *Y(t)* and *t* (Figure 1). The calculation of the correlation coefficient is very fast and can be completely automated for a large number of data, and implicitly it also provides a measure of the validity of the assumptions that lead to Equation 4.

This calculation can be updated day by day, and eventually, the extrapolation for *ln(N)* will converge to the correct number. As we will show in the results section, in many cases the end point can be predicted accurately at a very early stage.

The pseudo-code for data fitting is the following:

~~~
Read in csv date, Total Cases, Total Deaths for all the world
location
Correct errors, in the date
Main loop for each location
for line_end to End {
     for line_start 10 to line_end-10{
         step lnN from lnN1 to lnN2 by dlN{
              x=day; y = ln(N)-ln(X(t))
              CC = correlation_coef(x,y)
              Find maximum CC
         } if best CC > threshold
         Keep line Y coordinates and the lnN values
     }
}
~~~

For each line_end, collect the predicted *N* values and histogram them to find the most common value that is then taken as the prediction for that particular line-end value. We are well aware that this method can be improved in many ways some of which we are currently exploring.

### Data Smoothing

All data is smoothed using the LOWESS method (locally weighted scatter-plot smoothing) developed by W. S. Cleveland at Bell Labs in 1988 (Cleveland 1988). We use the original FORTRAN code written in Ratfor (Ratfor 1976) (https://www.netlib.org/go/lowess) and converted to Mortran (Mortran 1975). The parameter *F* (the fraction of points used to compute each fitted value) is set to 0.05, 0.07, 0.1, 0.12 and 0.14 for SMO1 to SMO5, respectively. In addition, the smoothed output Y-axis values for SMO4 and SMO5 are smoothed a second time using *F*=0.1. Smoothing is only applied to the total counts of cases and deaths. Well-aware of the distortions that smoothing can cause, we made sure that the smoothing did not introduce false features at the start or end of the time series. We also made sure that the smoothing did not move the location of the peaks as shown in **Fig. S3**. We also test the root-mean-square value of the change in total values caused by the five different levels of smoothing. When we do this for locations with more than 60 deaths and for locations with more than 1000 cases we find that the % RMS error average values are between 0.4% and 1.2% for *F* ranging from 0.05 to 0.14.

One problem when using smoothed data to test prediction, is that smoothing uses future data points that would not have been available on the day the prediction would have been made. We allow for this in estimating when new cases and deaths peak by taking the effective peak date for completed situations as half way between the actual peak date found in the smoothed data and the date at which the level has dropped past the peak to half peak height. We also generally avoid using smoothed data.

## RESULTS

### What To Expect From Simple Mathematical Functions

The most important result of this study is that the Gompertz function can be transformed into a straight line provided one knows the final plateau value of total counts of either cases or deaths, denoted here as *N*. This is shown in **Fig. 1** and provides the basic method we use to fit the observed data. Namely, vary the value of *N* to make the transformed Gompertz function *Y(t)* into a straight line and then derive parameters from the fit. Although this result is asymptotically true for a broad class of growth functions, we find that the simple three parameter Gompertz growth function fits the trajectory of actual COVID-19 outbreaks very well (Fig. 2). Specifically, the logarithm of the slope of ln*(X(t))* (called *H(t*)) decreases linearly with time meaning that the exponential growth rate (the slope of ln*(X(t))*) is never constant so that growth is never exponential. This linear decrease of *H(t)* is not true for all growth functions: specifically, the sigmoid function starts with pure exponential growth (**Fig. 2, c-d**). We find this linear decrease of ln*H(t)* is in fact a universal property of all outbreaks (Fig. 3) justifying the broad use of the Gompertz function here.

**Figure 2.**
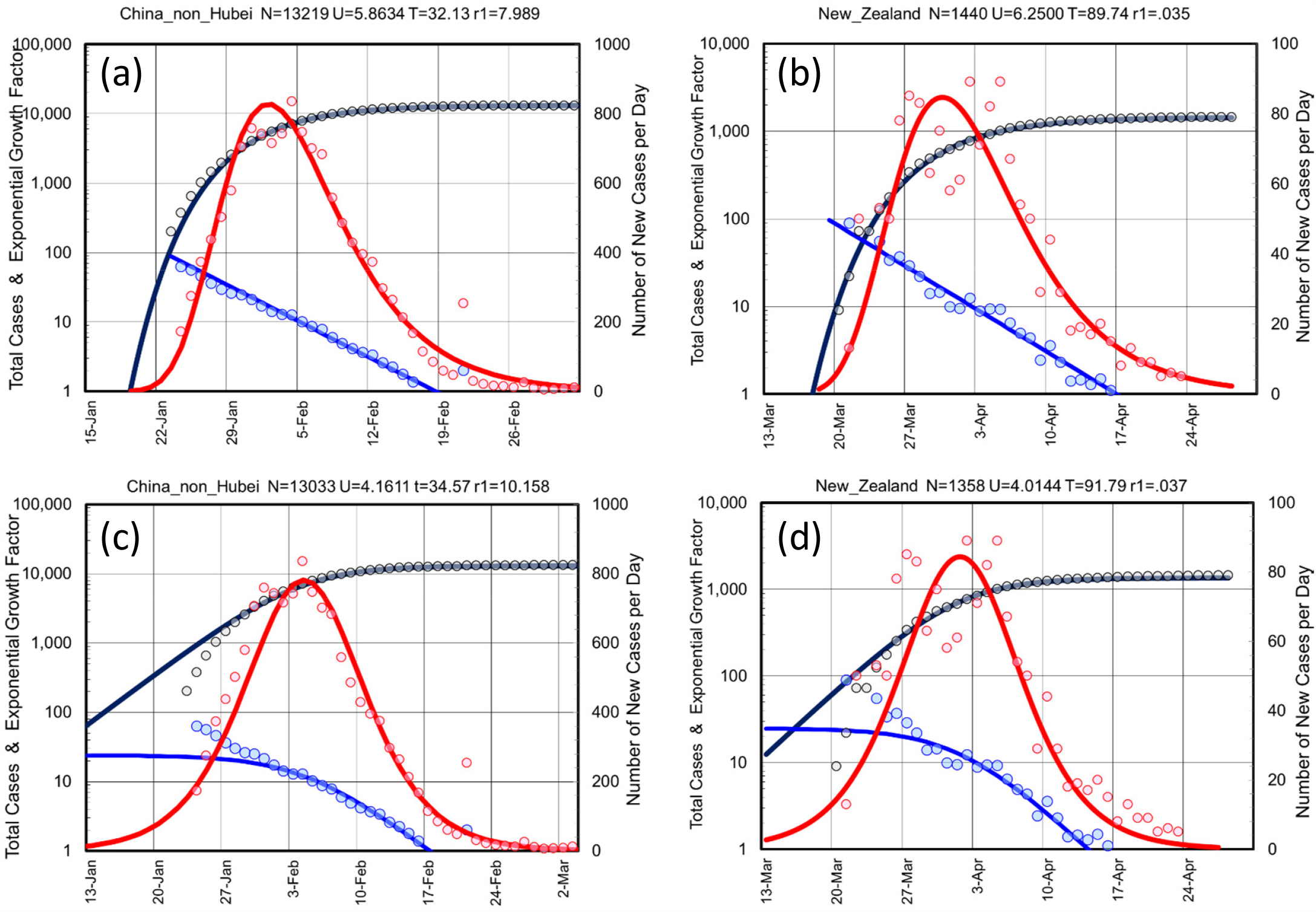
Showing How Early Raw Data Analysis Shows Non-Exponential Growth. Showing that the data from two outbreaks far apart in both space and time are almost indistinguishable. The raw data shows as colored circled of two well-controlled outbreaks in China, non_Hubei (all China except for Hubei Province) shown panels (a) & (c) and in New Zealand panels (b) & (d) are essentially identical. The fits for the data (solid lines) as also very similar except for the maximum plateau value of confirmed cases *N*=13,219 & 1,500, respectively) and the mid-point date in number of days from 23 January 2020, *T*=32.13 & 90.50. The *U* parameter is also very similar at *U*=5.87 & 5.88 days, respectively. Use of the Sigmoid Function in panels (c) & (d) give a fit that is less good that that obtained with the Gompertz Function in panels (a) & (b). This is shown by higher fit residuals (10.158 vs. 7.989 and 0.037 vs 0.035). More importantly, when compared to the Gompertz function, the Sigmoid function is less able to capture the behavior at the start of the outbreak. Following our four-panel graphs, we plot the Total Number of Cases (black line for *X(t)*, on left-hand y-axis, which is a log-scale), the number of New Cases (red line for *X(t)-X(t-1)*, right hand y-axis, which is a linear scale), and Gradient of log Total Cases (blue line for *H(t) = ln(X(t)) – ln(X(t-1)) = ln(X(t)/X(t-1))* on the left-hand y-axis, log scale). Note that for both the real data and the Gompertz function, ln[ln*[(X(t)/X(t-1)*)]] is a linear function of time, *t*.

**Figure 3.**
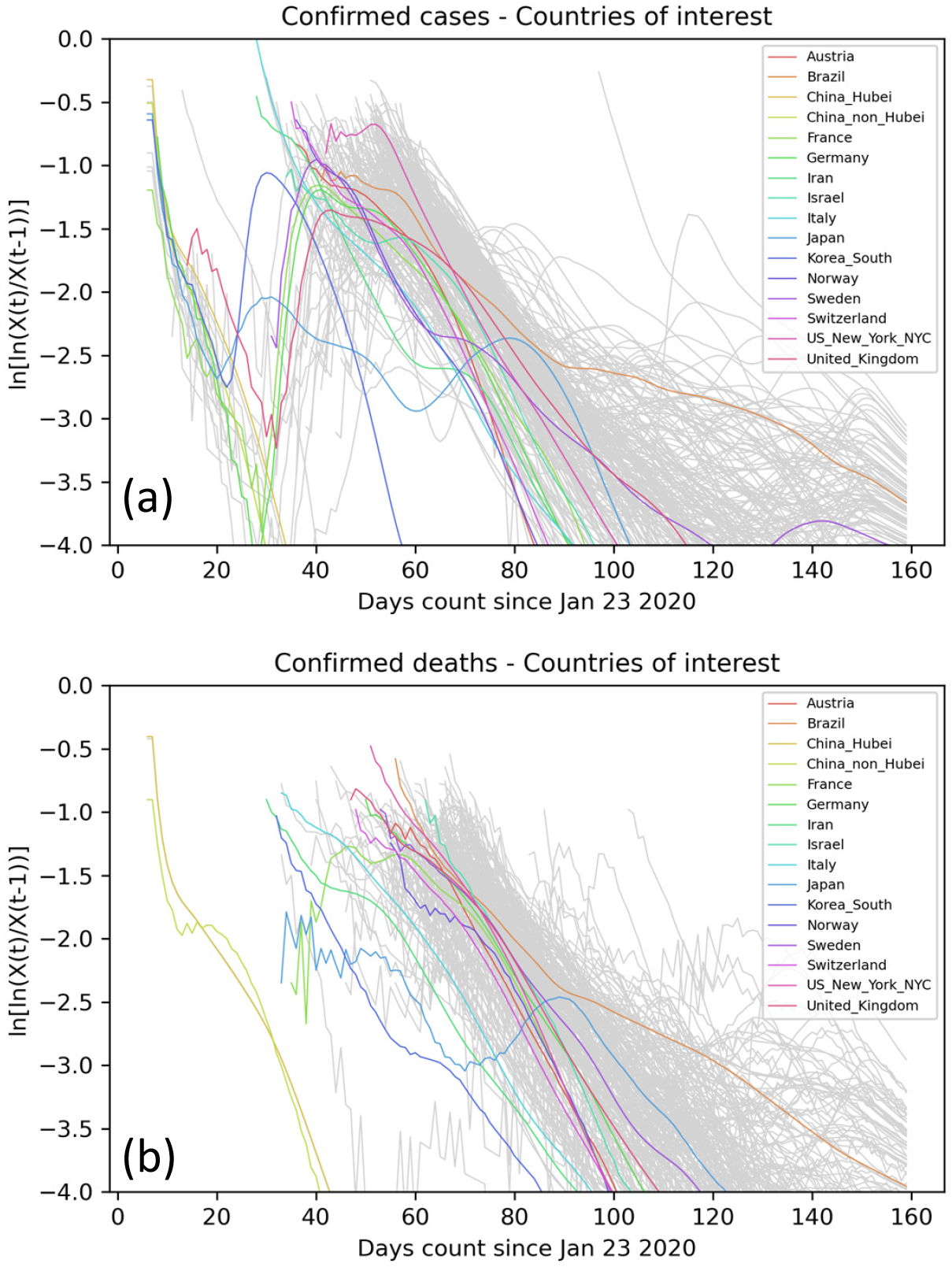
Value *ln[H(t)]=ln[ln[X(t)/X(t-1)]]* Deceases Linearly as Expected for Gompertz Function. (data is smoothed with SMO5 as difference of small numbers) Showing the trajectory of ln[*H(t)*] or ln[ln[*X(t)/X(t-1*)]] for all selected locations with more than 50 deaths. From **Fig. 2**, ln[*H(t)*] is expected to decrease linearly for the Gompertz function. As *H(t)=*ln[*X(t)*] – ln[*X(t-1*)] is the difference of two numbers, it is subject to a high level of noise. For this reason, we smooth the *X(t)* using SMO5 LOWESS smoothing. Panel (a) shows the trajectories of ln[*H(t)*] for cases. Panel (b) shows ln[*H(t)*] for deaths. As there are often relatively low numbers of deaths, the trajectories for deaths are still noisy even after smoothing (NB. The noise in some highlighted locations is unexpectedly high and warrants further investigation).

### Classification of World COVID-19 Outbreaks

Table 1 lists those countries (89 in all) or regions (Italy, US & Canada, 147 in all) with more than 50 deaths or 1000 cases. The outbreaks have been classified by our completeness code that is based on the peaking of the number of new daily cases or new daily deaths. (See **Table 1** for explanation for explanation of the completeness code).

**Table 1:**
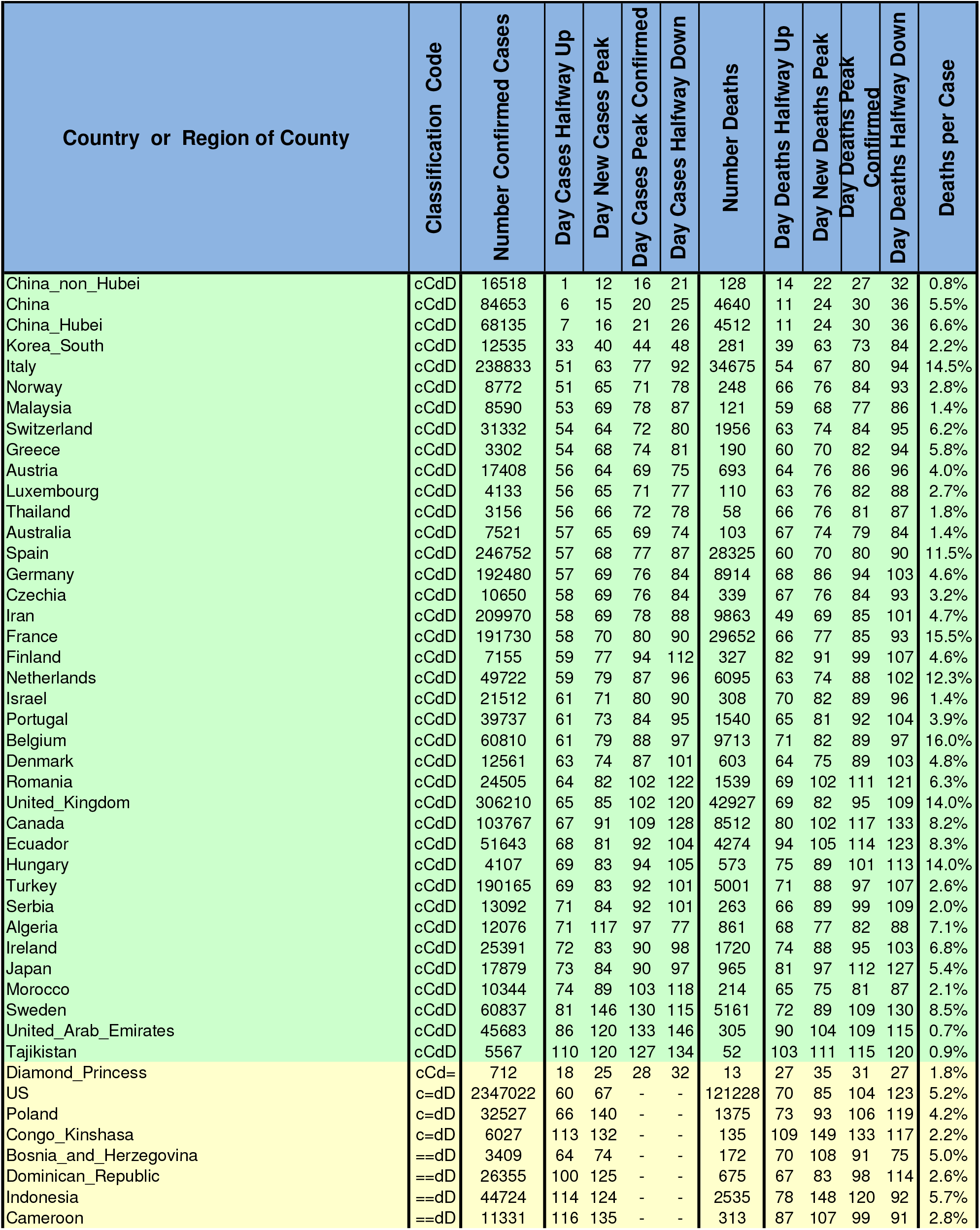
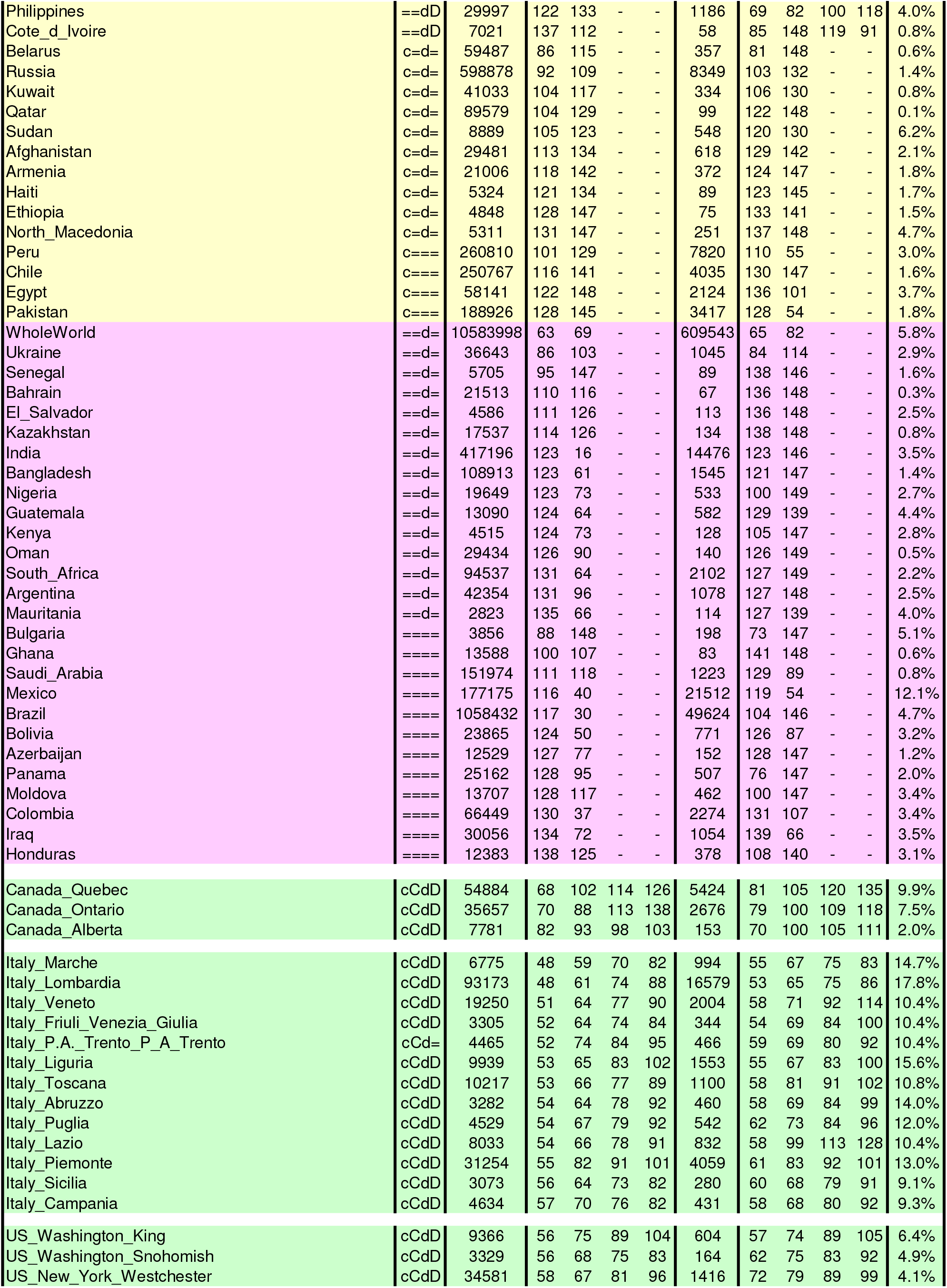
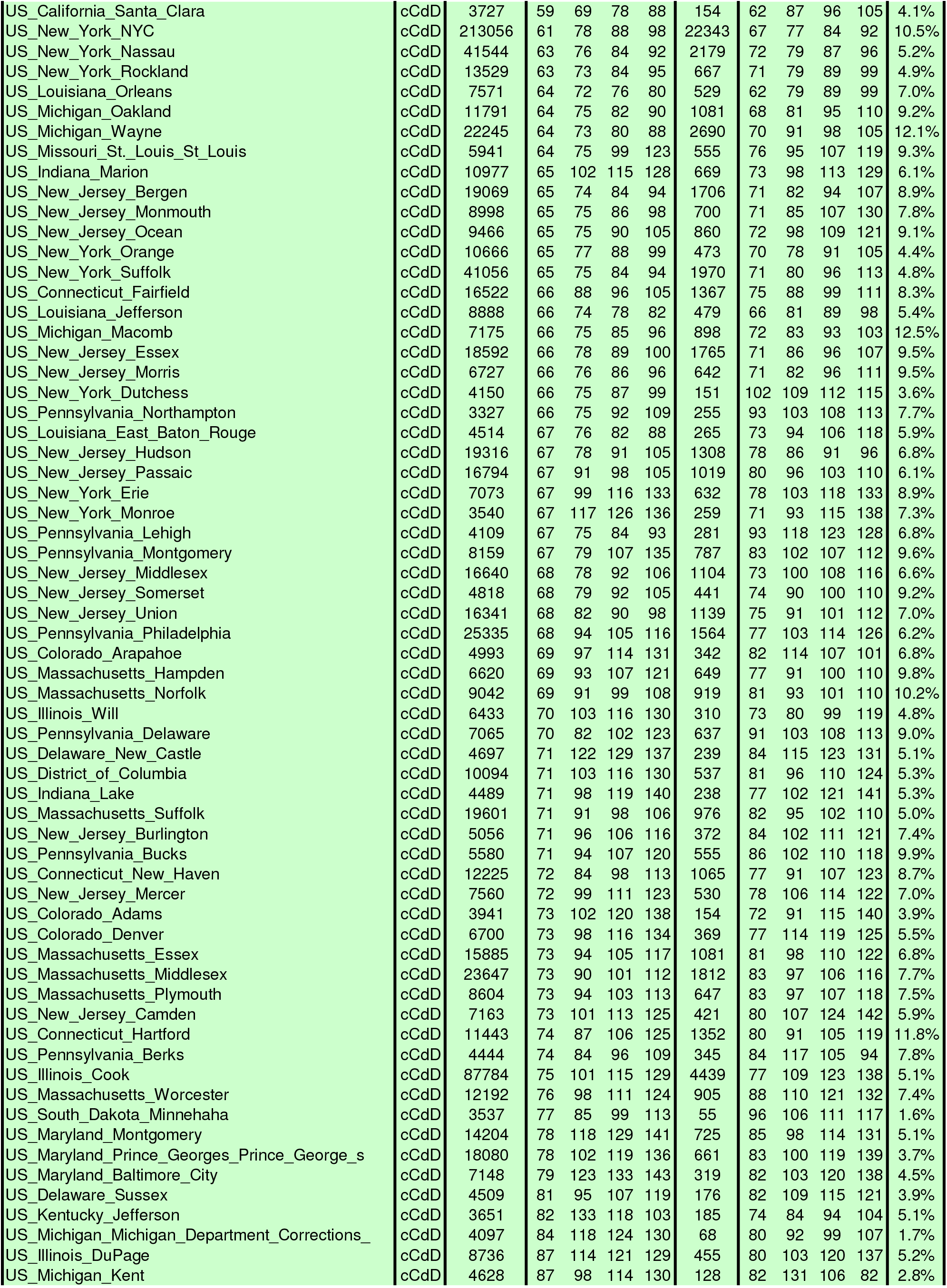
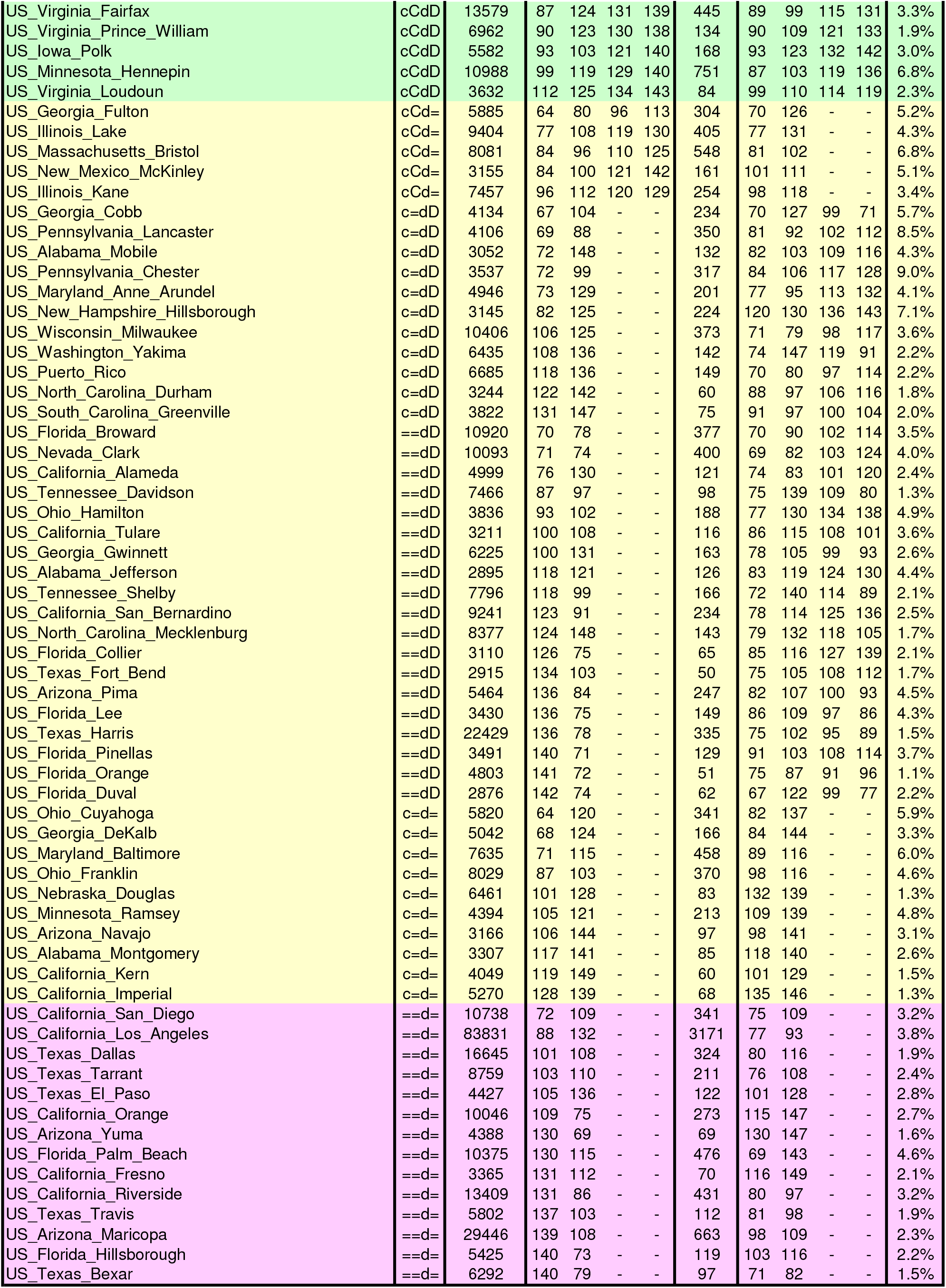
Classification of Selected Locations (LOWESS smoothing SMO5 used to find peaks). KEY: ‘c’ means New Cases peaked, ‘C’ means at least halfway down this peak, ‘d’, ‘D’ are same for New Deaths. We also give Day of Peak, Day Halfway Up, and Day Halfway Down. Day Peak Confirmed is defined as (Day of Peak + Day Halfway Down)/2. Showing the classification scheme we use for all worldwide outbreaks. The Classification Code consist of four symbols, two for Cases and two for Deaths that are initially set to ‘=‘. Position 1 is ‘c’ if New Cases per Day have reached a maximum and are dropping; position 2 is set to ‘C” if New cases per Day have dropped to below half the maximum values; positions 3 & 4 are set to ‘d’ and ‘D’ when new deaths per day have reached a maximum or have dropped to half their maximum value. The determination of peaking is made using heavily smoothed data (SMO5) (see text).

### Fitting With a Straight Line

**Fig 4 (a)** shows the function *Y(t)* for deaths in the many different locations (countries or regions of countries) which have reached a plateau, and for which the prediction of the final *N* is stable. It is evident that for all these locations the data generally follows a linear relationship thereby justifying *a posteriori* our working hypothesis. This observation is confirmed by the fact that the correlation coefficient with time of *Y(t)* is close to 1 for the vast majority of the locations we examined. We also note that the time constants *U* (i.e. the inverse slope of the lines or the time-constant of decay) are very similar to each other, indicating the existence of universal properties in virus diffusion that are largely independent of the country.

**Figure 4.**
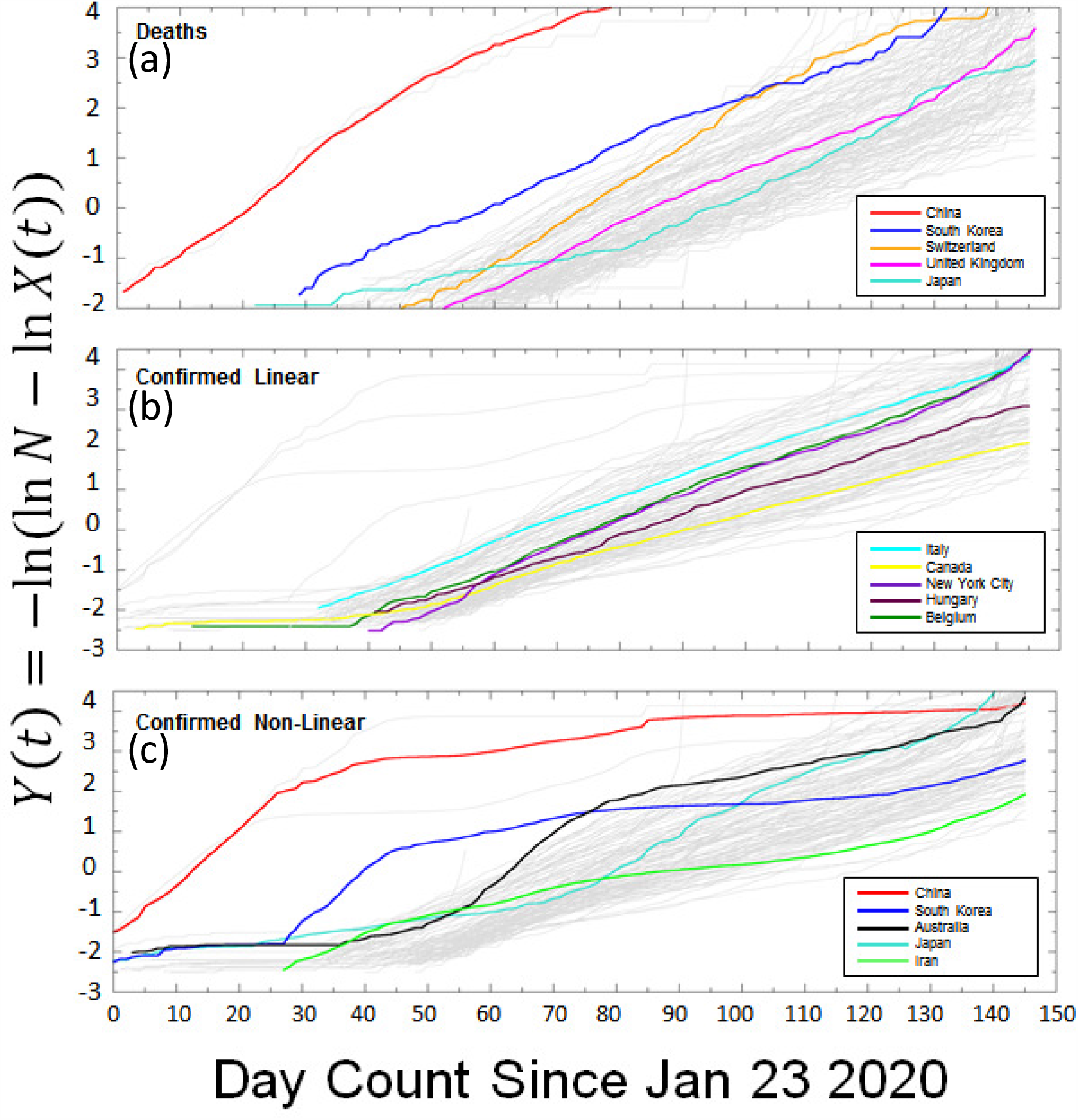
Function *Y(t)=ln[ln[N/X(t)]]* Give Straight Lines for Raw Unsmoothed Data. Showing the trajectory of *Y(t))* or –*ln(ln(N/X(t)) = −ln(ln(N)-ln(X(t)))* for all selected locations with more than 50 deaths. From **Fig. 1**, *Y(t))* is expected to decrease linearly for the Gompertz function and, for more general growth functions in the limit of large *t* (see **Methods**). In panel (a) we show the raw data for *Y(t)* for deaths in a selection of more than 130 locations (thin gray lines), and emphasize 5 representative ones with a thicker line. For all such selected locations, *Y(t)* is well approximated by straight lines with a very similar slope. Panels (b) shows *Y(t)* for confirmed cases in 119 locations. In panel B we emphasize locations for which the function *Y(t)* is again well approximated by a straight line, while in Panel (c) we show some locations for which this is not true anymore. This is expected if multiple outbreak of comparable intensity happens in a country, or if there is change in the dynamics of infections or the way cases are counted.

When considering confirmed cases (**Fig. 4**), we observe more diverse behavior in the time course of *Y(t*). While for some countries the linear relationship still holds (**Fig 4 (b)**), in other countries we notice deviation from linearity (**Fig. 4 (c)**), which could indicate the existence of multiple outbreaks, or could reflect a change in the method of counting cases.

By fitting *Y(t)* to a single straight line, we can average multiple outbreaks into a single major outbreak which will follow a Gompertz distribution, where the parameters *U* and *T* are the slope and the x-axis intercept at *Y(t)*=0, of *Y(t)*. This approach allows us to obtain a uniform description for every time series *X(t)* of cases and deaths in different parts of the world, but with loss of details for locations that do not follow a simple linear relationship.

While *a posteriori* fits describe the raw data well, extrapolations of the final plateau before a given day are still subject to large fluctuations, due to the (double) exponential nature of Gompertz law. In other words, when *X(t)* is small compared to *N*, the fitting line varies approximately like ln(ln*(N*));.even large variation of *N* barely affects the quality of the fit. Vice versa, when *X(t)* approaches *N, Y(t)* becomes more and more sensitive to the correctness of the predicted value of *N* (**Fig. S2**) The consensus predicted value of *N* converges to a plateau value with time, and then it is followed by real data with some delay. This allows us to discriminate between locations in which confirmed cases (or deaths) have reached or are near to reach the plateau, and locations for which it is still impossible to predict the plateau.

### A closer Look at Specific Locations

While COVID-19 trajectories share many properties, each outbreak has its own features, which affect our ability to forecast the outcome in terms of the plateau value *N* for both cases and deaths. These features are best appreciated using two types of graphs, the Four-Panel graph and the Best Line Prediction graph described carefully in **Fig. 5**, which shows these two graphs for Germany a large but well-behaved outbreak. The top panel of **Fig. 5(a)** shows that for smoothed data there are single peaks of new cases and new deaths, with the new deaths peaking 11 days after the new cases. This is almost exactly what we observed for the smaller and much earlier outbreak in China, non_Hubei, where deaths were most likely to occur 10 days after a case was confirmed (**Levitt, 2020f**); this suggests that this interval may be connected to the natural progression of the disease in well-managed scenarios. The same delay between cases and deaths is also seen (as it should be) in the second panel. The third panel shows the characteristic curvature recognized since our 14-Mar-20 analysis (Levitt 2020b). Together with the forth panel, it also reveals a small initial outbreak that started on 24-Jan-20, was contained and then followed by a much larger outbreak that started two weeks later and became clearly seen after another two weeks. The Best Line Prediction (BLP) graphs for Germany show in **Fig. 5 (b)** that from 1-Apr-20, the plateau value of total cases would have been well-predicted. For deaths, **Fig. 5 (c)** shows the eventual plateau value could have been predicted accurately on 10-Apr-20. The blue dots on these two graphs show that the predicted plateau values vary wildly and a prediction can only be made because many straight-line fits give a similar consensus *N* value.

**Figure 5.**
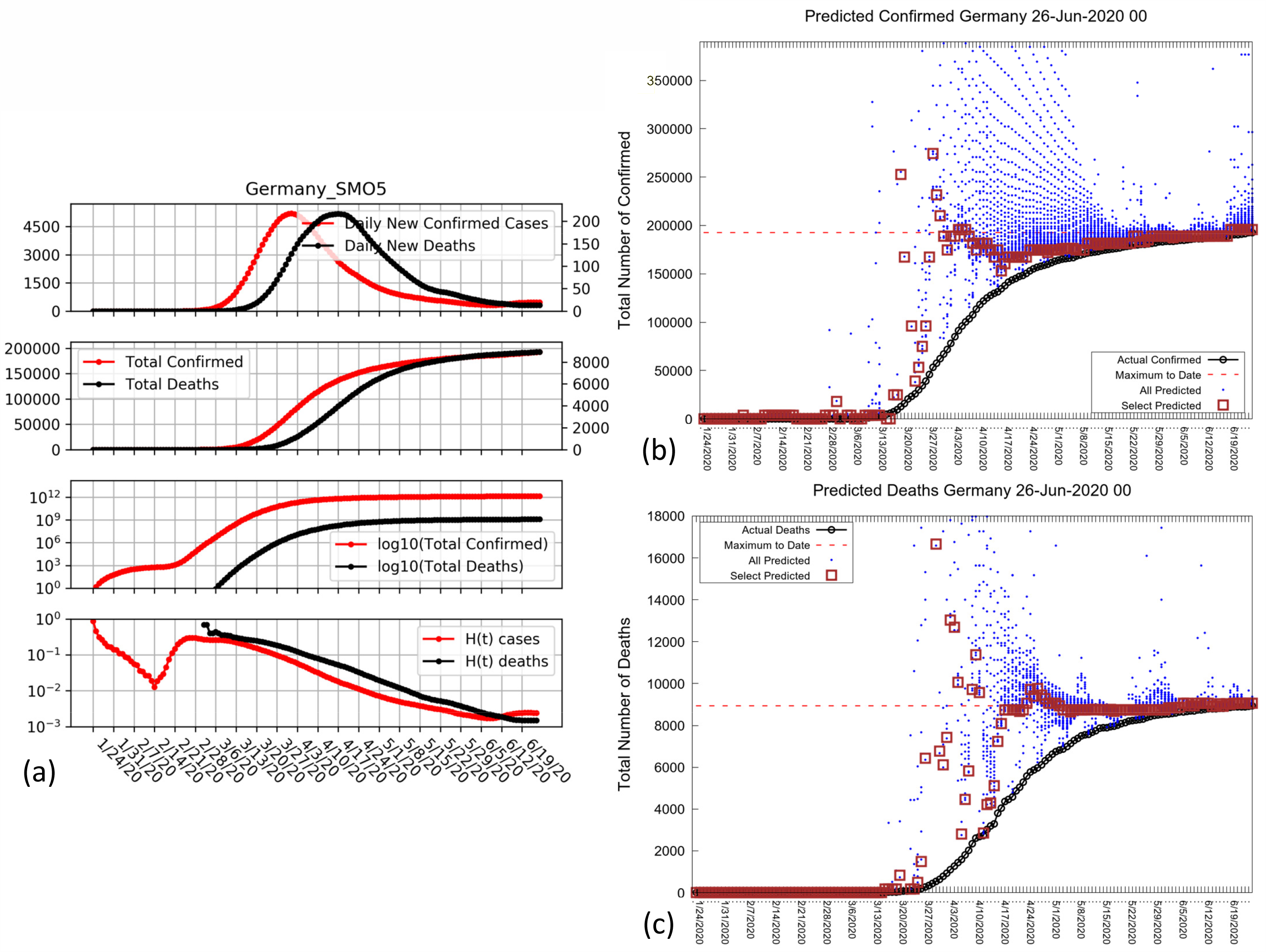
Showing Best Line and Smooth Peak Graphs for Germany. Showing for Germany the Four-Panel graph (left) and Best Line Prediction (BLP) graphs right. In the Four-Panel graph, which has been carefully refined since Feb. 2020 to show the most relevant data in an epidemic, the top panel is New Cases per Day (red, left y-axis) and New Deaths per Day (black, right y-axis) both normalized to the same height. The second panel is Total Cases and Deaths shown in their respective color and y-axis and also normalized. The third panel is these same totals plotted on a log_10_ scale (no need for normalization). The fourth panel is ln[(*X(t)/X(t-1)* plotted on a log_10_ scale (log of the fractional change used in our first analysis, Levitt 2020c). Here we consistently use log_e_ or ln for calculations as growth functions are defined in terms of the exponential, *e*; we use log_10_ to define logarithm y-axes as powers of ten are more familiar to us humans than are powers of *e*. In the BLP graph, Dates are plotted along the x-axis and Total Number of either Cases or Death along the y-axis. The actual trajectory of total data counts is plotted as heavy black circles and increases monotonically with time. The horizontal red dashed line marks the maximum total count number on the latest day included (date specified in the title). The blue dots are the candidate predicted *N* plateau values (the predicted final completion total count) shown at the Date value where they were made. Specifically, only data up to including this date can be used to find the ln(*N*) value that gives the best line. The brown squares enclose the actual predicted *N* value that is found by most of the predictions at that date (most overlapping blue dots are in the boxes).

In **Fig. 6** we show four other locations which have reached a plateau and for which the extrapolation has not changed significantly in the last few weeks. Although none of these locations are as clean as Germany (**Fig. 5**), one see that early predictions are unstable but converge to more realistic figures with time. **Fig. 6(a)** shows New York City to be well-behaved in terms the smoothed peaks of new cases and new deaths although deaths and cases seem to occur at the same time. This suggests a situation less under control than either China, non_Hubei or Germany. Nevertheless, the BLP graph shows that the final plateau value of *N* for deaths in New York City could have been predicted correctly on 10-Apr-20. The decay of *H(t)* shown in the bottom panel of the four-panel graph is very clean suggesting a single outbreak. **Fig. 6(b)** shows Sweden to have very badly formed peaks of new cases and new deaths and deaths seem to occur before cases, an impossibility likely due to decreased counting of cases as the epidemic proceeds. The BLP graph predicts a plateau value that is not constant although it does look as if total deaths will plateau at about 6,000. **Fig. 6(c)** shows Russia to also have a very extended peaks of new cases and deaths. The number of new cases peaked 7 weeks ago but new deaths remain high. Nevertheless, the BLP graph shows that the predicted plateau value of *N* for deaths is increasing more and more slowly and may well converge to a value of about 16,000, almost double the current number of deaths. **Fig. 6(c)** shows Mexico deaths to be increasing even more rapidly than Russia and at present it is impossible to predict the plateau.

**Figure 6.**
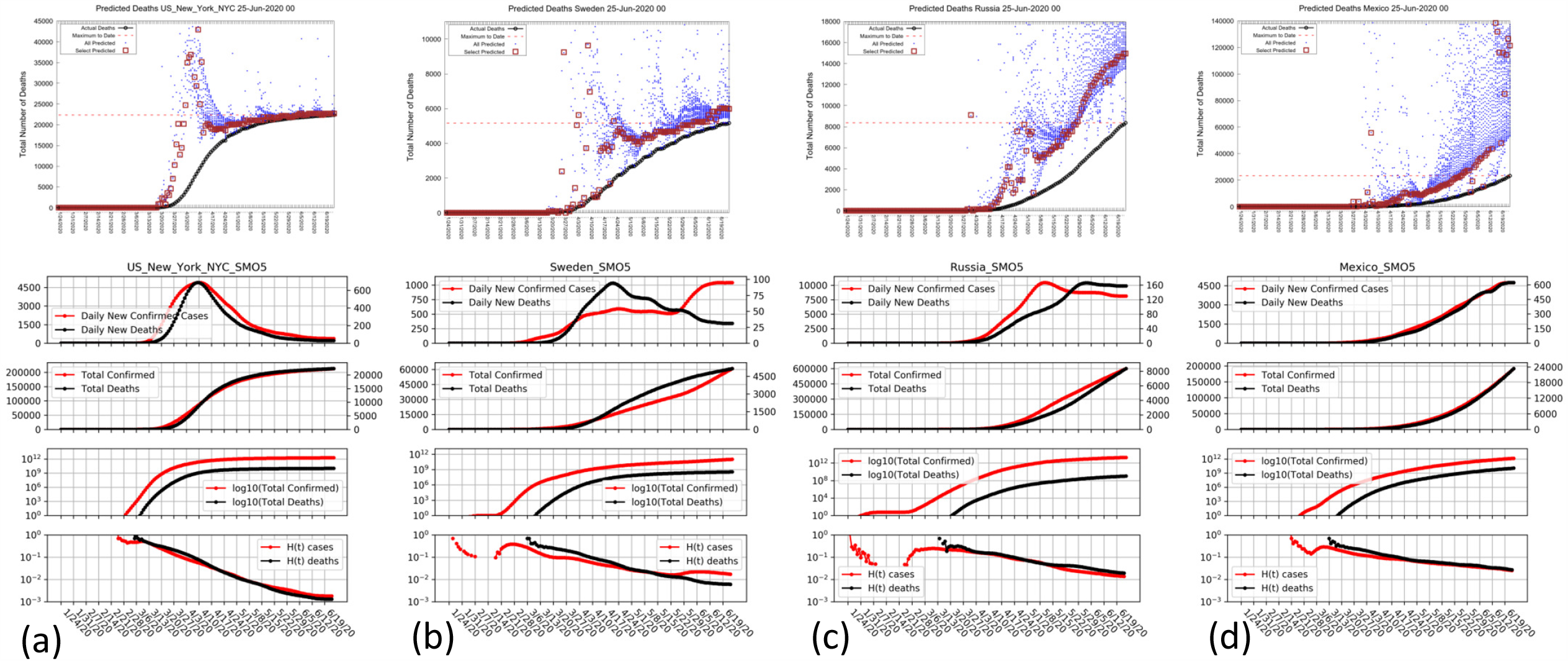
Plots showing Best Line and Smooth Peak Graphs for Selected Locations. Showing four locations, which behave differently because they are at different stages of their outbreak. (a) Deaths in New York City, which was the hardest hit location with more deaths per population than anywhere else. The smoothed data in the lower part of the Four-Panel graph shows clean peaks for Cases and Deaths and a linear descent *H(t)* on the log scale. The Best Line Prediction in the upper part shows that the plateau number of deaths was indicated as early as 4-Apr-20 and confirmed a week later. (b) Deaths in Sweden, which adopted very limited social distancing and no lockdown. The smoothed curve of new cases and new deaths remains elevated for much longer than in NYC although there is a very similar linear descent *H(t)* on the log scale. The BLP seems to edge up but a good prediction of the current plateau could have been made on 22-Apr-20. This is 10 days earlier than a prediction of Sweden peaking we made on Twitter on 2-May-20 (Levitt-Twitter) showing the power of the BLP method that we did not have back then. (c) Confirmed Cases Russia are growing rapidly, although the number of new cases per day peaked on 8-May-20 they remain stable at a high level. The BLP method tentatively predicts a plateau *N* value of about 700,000 cases in Russia. (d) Deaths in Mexico are still far from any clear plateau value..

In **Table 2**, we compare the predictive power on the Best Line Prediction (BLP) with that of the Peak Detection Method (PDM). Checking all the converged locations where the current value is expected to close to the expected plateau value of *N* shows that the BLP is significantly better than the PDM. Both methods seem to be able to make their predictions at about the same time (on average, the PDM predicts two days earlier than the BLP based on our assumed value for the peak confirmation date.

**Table 2:**
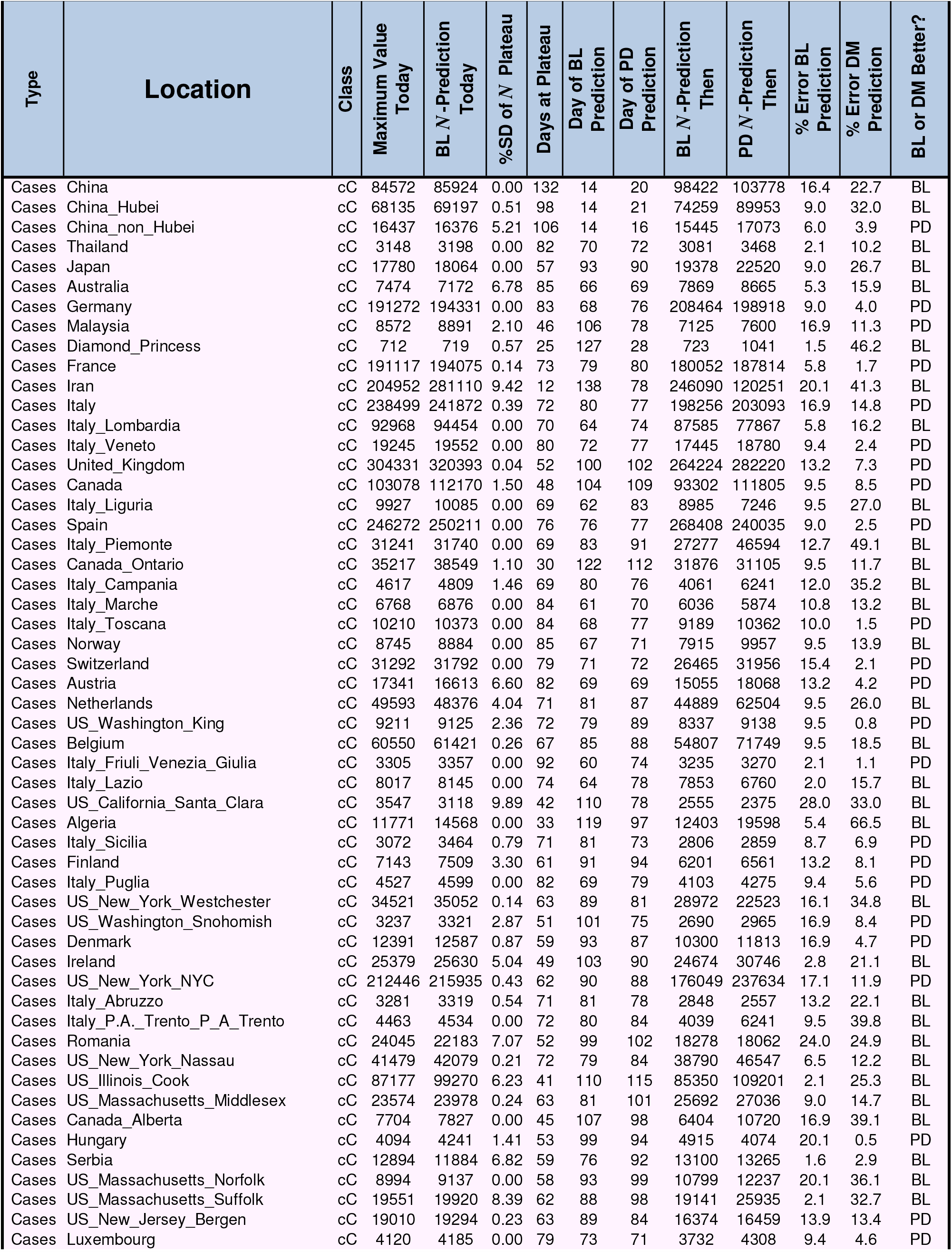
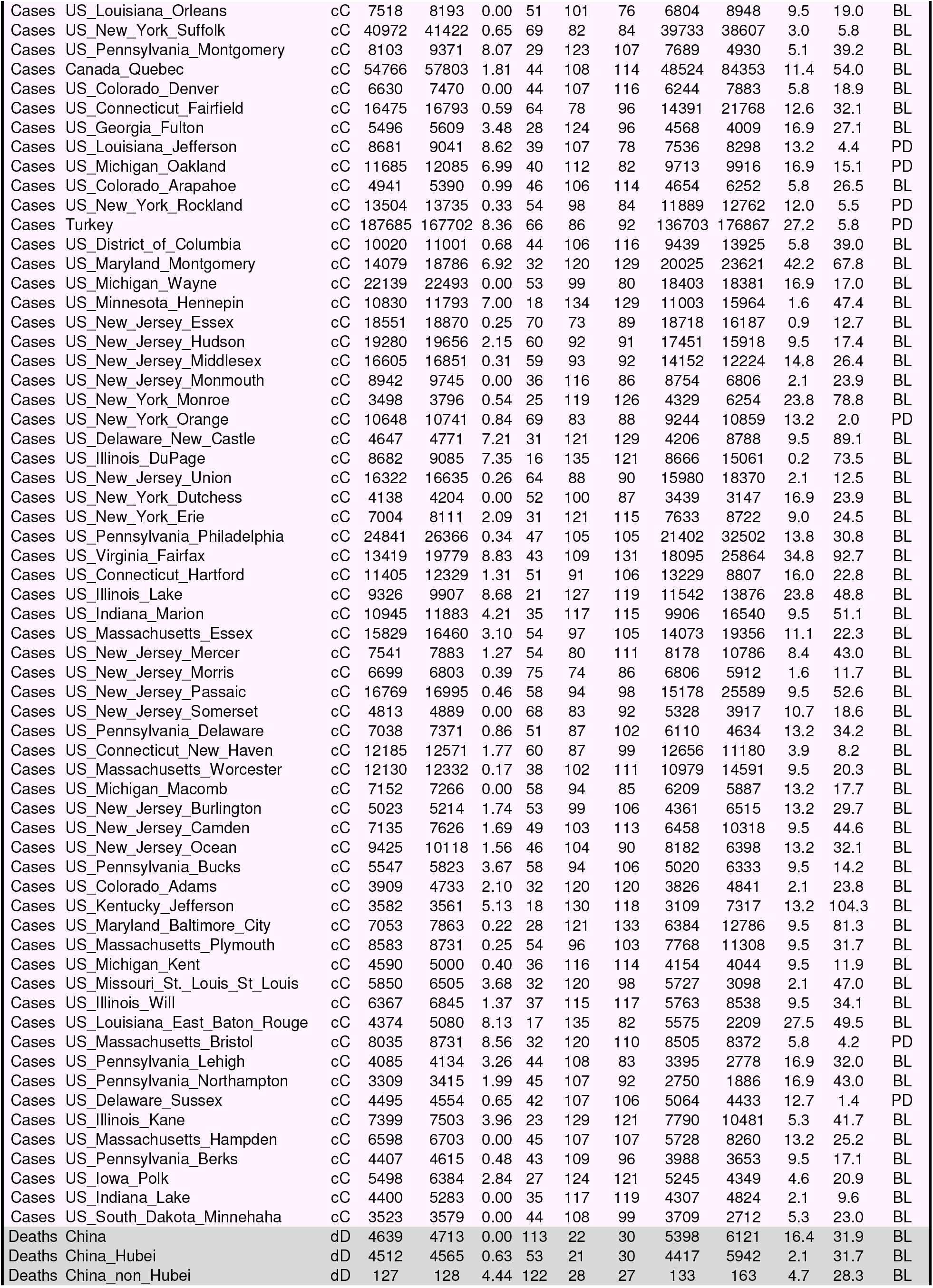
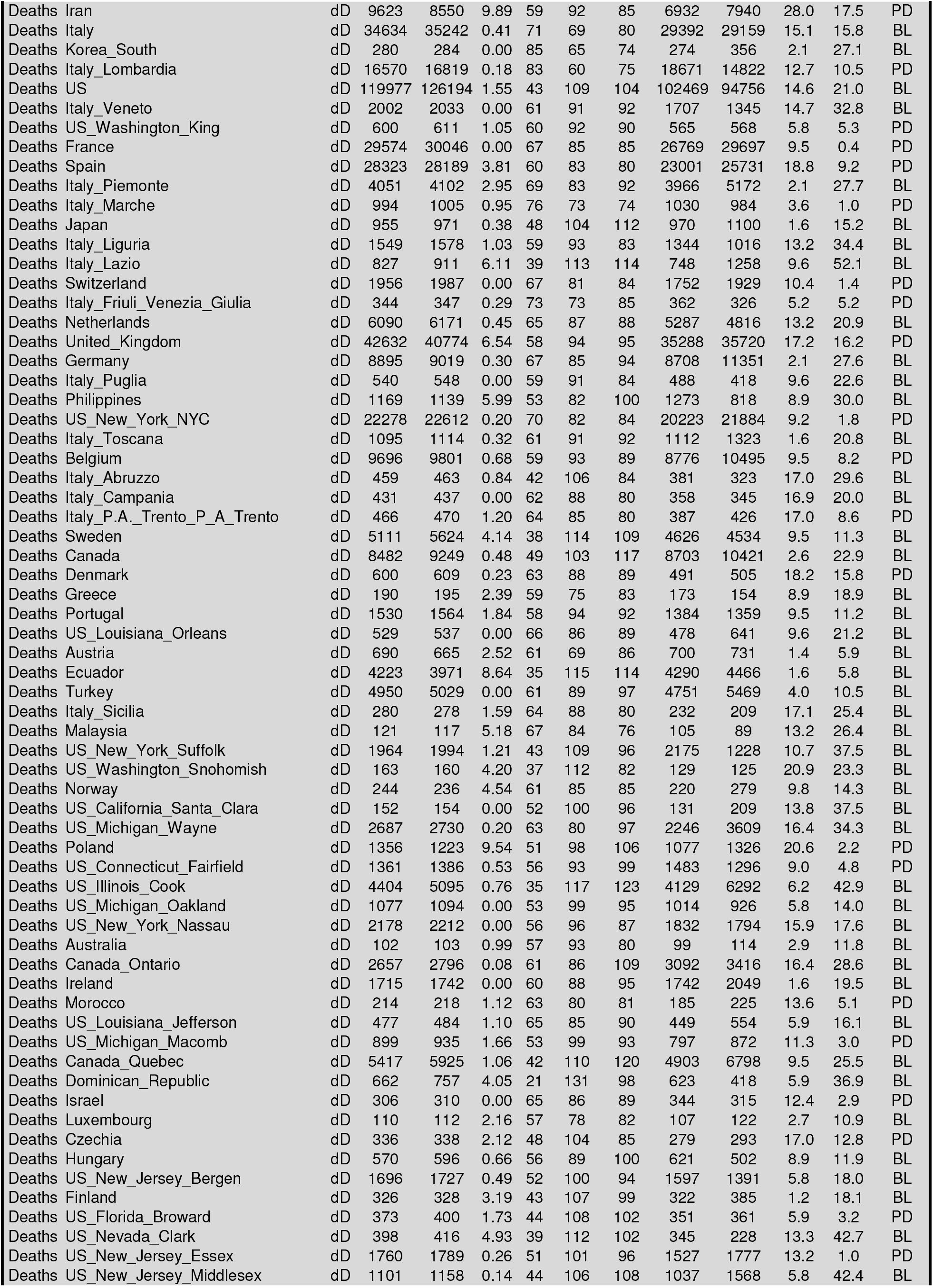
Comparing Best Line (BL) and Peak Detection (PD) Prediction of Plateau *N* Value. Comparing Best Line Prediction (BLP) and Peak Detection Method (PDM) for Prediction of Plateau *N* Value. The plateau *N* value predicted by the Best Line method is significantly more accurate than that predicted by the Peak Detection method. This can be measured by the Percent Error of the Prediction defined as 100*(Predicted_Plateau_Value - Value_Now)/(Value_Now). For the BLP method this number averages 11% for cases prediction and 9.5% for deaths prediction, whereas the corresponding values for the PDM are more than double at 25.3% and 23.7%, respectively. Another way to measure the advantage of BLP over PDM is to count for different locations how often BLP does better than PDM. Here BLP is better than PDM in 74% of the locations for cases and in 73% of the locations for deaths.

In **Table 3** we look at the most active locations to identify cases where prediction of outcome could have significant impact. For this we use two criteria: First, that the forecast be reliable in that the plateau is stable in terms of its slope, its percent standard deviation and at least seven days at this plateau value. Second, that the forecast plateau is a significant increase over the current level.

**Table 3.** Forecasts of Plateau *N* Ordered by Size and Certainty (green shading more certain but may involve small increases to plateau so less important). The locations here are not converged (their classification code is not ‘cCdD’). Rather than look at the individual Best Line graphs manually, we do line fitting to the predicted plateau *N* working back from today. Key parameters are the slope of the line th**r**ough the plateau values, which should **be** small, the Percentage Standard Deviation of the Plateau value (%SD) and the number of days with a plateau prediction within 20% of the predicted value.

| Cases or Deaths | Location | Total Number Now | Predicted Plateau *N* Now | %SD of Plateau | Difference Now to to Plateau | %Difference | Number Days at Plateau | Slope of Plateau with Day |
| --- | --- | --- | --- | --- | --- | --- | --- | --- |
| Cases | Peru | 257,447 | 478,018 | 1.3 | 220,571 | 85.7 | 22 | 19 |
| Deaths | Brazil | 51,271 | 97,987 | 3.6 | 46,716 | 91.1 | 16 | 42 |
| Cases | Belarus | 59,023 | 82,618 | 5.7 | 23,595 | 40.0 | 22 | 158 |
| Cases | Indonesia | 46,845 | 68,939 | 5.0 | 22,094 | 47.2 | 8 | 395 |
| Cases | Afghanistan | 29,157 | 45,900 | 7.4 | 16,743 | 57.4 | 10 | 215 |
| Cases | United_Arab_Emirates | 45,303 | 57,910 | 6.8 | 12,607 | 27.8 | 31 | 25 |
| Cases | Philippines | 30,682 | 41,752 | 2.6 | 11,070 | 36.1 | 12 | 259 |
| Cases | Kuwait | 40,291 | 46,889 | 3.4 | 6,598 | 16.4 | 31 | 233 |
| Deaths | Peru | 8,223 | 14,544 | 8.8 | 6,321 | 76.9 | 24 | 144 |
| Cases | US_Tennessee_Davidson | 7,716 | 13,730 | 5.3 | 6,014 | 77.9 | 8 | 423 |
| Cases | Cameroon | 12,041 | 17,830 | 9.6 | 5,789 | 48.1 | 14 | 480 |
| Deaths | Russia | 8,196 | 13,573 | 6.6 | 5,377 | 65.6 | 20 | 151 |
| Cases | El_Salvador | 4,808 | 9,177 | 9.2 | 4,369 | 90.9 | 8 | 492 |
| Cases | Dominican_Republic | 27,370 | 31,447 | 6.0 | 4,077 | 14.9 | 14 | 637 |
| Cases | US_Washington_Yakima | 6,326 | 10,266 | 3.4 | 3,940 | 62.3 | 6 | 167 |
| Cases | Sudan | 8,698 | 12,425 | 2.8 | 3,727 | 42.8 | 20 | 174 |
| Deaths | Bangladesh | 1,502 | 4,870 | 8.8 | 3,368 | 224.2 | 7 | 212 |
| Cases | Moldova | 14,363 | 17,669 | 8.5 | 3,306 | 23.0 | 10 | 508 |
| Cases | US_Texas_El_Paso | 4,553 | 7,645 | 7.6 | 3,092 | 67.9 | 12 | 285 |
| Cases | US_Maryland_Baltimore | 7,585 | 10,177 | 8.9 | 2,592 | 34.2 | 20 | 0 |
| Cases | US_Maryland_Anne_Arundel | 4,916 | 7,175 | 0.1 | 2,259 | 46.0 | 6 | 1 |
| Cases | US_Nebraska_Douglas | 6,386 | 8,611 | 0.0 | 2,225 | 34.8 | 13 | 0 |
| Cases | US_Ohio_Franklin | 7,915 | 9,941 | 3.3 | 2,026 | 25.6 | 18 | 97 |
| Cases | US_California_Orange | 10,595 | 12,609 | 9.8 | 2,014 | 19.0 | 16 | 301 |
| Cases | Ukraine | 38,056 | 39,656 | 9.6 | 1,600 | 4.2 | 19 | 754 |
| Cases | Haiti | 5,211 | 6,480 | 3.2 | 1,269 | 24.4 | 8 | 38 |
| Deaths | Indonesia | 2,500 | 3,449 | 7.1 | 949 | 38.0 | 9 | 16 |
| Cases | US_Georgia_Gwinnett | 6,407 | 7,219 | 8.9 | 812 | 12.7 | 16 | 169 |
| Cases | US_California_San_Diego | 11,096 | 11,875 | 7.4 | 779 | 7.0 | 23 | 131 |
| Cases | US_Ohio_Cuyahoga | 5,734 | 6,450 | 7.6 | 716 | 12.5 | 8 | 184 |
| Deaths | Argentina | 1,043 | 1,581 | 9.7 | 538 | 51.6 | 9 | 59 |
| Cases | US_Georgia_Cobb | 3,969 | 4,488 | 6.2 | 519 | 13.1 | 28 | 0 |
| Deaths | Ukraine | 1,022 | 1,539 | 4.3 | 517 | 50.6 | 9 | 31 |
| Cases | US_Puerto_Rico | 6,564 | 7,056 | 7.2 | 492 | 7.5 | 12 | 152 |
| Cases | US_Pennsylvania_Chester | 3,513 | 3,975 | 10.0 | 462 | 13.2 | 34 | 19 |
| Deaths | US_California_Los_Angeles | 3,137 | 3,506 | 2.7 | 369 | 11.8 | 31 | 25 |
| Cases | US_Minnesota_Ramsey | 4,352 | 4,675 | 7.2 | 323 | 7.4 | 23 | 34 |
| Deaths | Nigeria | 525 | 840 | 6.2 | 315 | 60.0 | 7 | 29 |
| Cases | US_Wisconsin_Milwaukee | 10,355 | 10,668 | 5.5 | 313 | 3.0 | 26 | 89 |
| Deaths | Armenia | 360 | 666 | 6.7 | 306 | 85.0 | 9 | 19 |
| Deaths | US_Florida_Palm_Beach | 468 | 747 | 7.0 | 279 | 59.6 | 10 | 14 |
| Deaths | US_Arizona_Maricopa | 634 | 776 | 8.4 | 142 | 22.4 | 29 | 9 |
| Deaths | US_Texas_Dallas | 317 | 434 | 3.0 | 117 | 36.9 | 32 | 0 |
| Deaths | US_Massachusetts_Bristol | 546 | 663 | 1.4 | 117 | 21.4 | 26 | 0 |
| Deaths | US_Ohio_Cuyahoga | 339 | 456 | 3.3 | 117 | 34.5 | 14 | 1 |
| Deaths | US_Maryland_Baltimore | 455 | 543 | 1.4 | 88 | 19.3 | 34 | 1 |
| Deaths | Sudan | 533 | 615 | 7.3 | 82 | 15.4 | 10 | 8 |
| Deaths | US_California_Riverside | 424 | 498 | 1.4 | 74 | 17.5 | 34 | 0 |
| Deaths | US_Texas_El_Paso | 120 | 190 | 7.2 | 70 | 58.3 | 6 | 5 |
| Deaths | US_Illinois_Kane | 252 | 319 | 8.1 | 67 | 26.6 | 21 | 0 |
| Deaths | US_Texas_Tarrant | 208 | 274 | 2.0 | 66 | 31.7 | 18 | 0 |
| Deaths | Kuwait | 330 | 396 | 3.2 | 66 | 20.0 | 18 | 1 |
| Deaths | US_California_San_Diego | 338 | 397 | 5.9 | 59 | 17.5 | 36 | 0 |
| Deaths | Bulgaria | 207 | 265 | 9.8 | 58 | 28.0 | 16 | 0 |
| Deaths | US_Ohio_Franklin | 358 | 408 | 2.8 | 50 | 14.0 | 17 | 3 |
| Deaths | US_Georgia_DeKalb | 165 | 205 | 8.5 | 40 | 24.2 | 32 | 1 |
| Deaths | US_Utah_Salt_Lake | 102 | 140 | 4.0 | 38 | 37.3 | 10 | 2 |
| Deaths | US_Colorado_Adams | 153 | 186 | 4.0 | 33 | 21.6 | 25 | 0 |
| Deaths | US_Illinois_Lake | 401 | 433 | 6.5 | 32 | 8.0 | 24 | 0 |
| Deaths | US_New_Jersey_Mercer | 524 | 556 | 8.3 | 32 | 6.1 | 46 | 0 |
| Deaths | US_California_Kern | 60 | 84 | 1.5 | 24 | 40.0 | 22 | 0 |
| Deaths | US_Nevada_Clark | 400 | 419 | 6.2 | 19 | 4.8 | 39 | 0 |
| Deaths | US_Maryland_Anne_Arundel | 200 | 217 | 1.9 | 17 | 8.5 | 41 | 0 |
| Deaths | US_Virginia_Loudoun | 84 | 100 | 3.0 | 16 | 19.0 | 6 | 1 |
| Deaths | US_Nebraska_Douglas | 80 | 94 | 6.0 | 14 | 17.5 | 6 | 1 |
| Deaths | US_Florida_Hillsborough | 115 | 128 | 9.8 | 13 | 11.3 | 15 | 3 |
| Deaths | Panama | 521 | 529 | 9.7 | 8 | 1.5 | 17 | 10 |
| Deaths | US_Puerto_Rico | 149 | 156 | 0.4 | 7 | 4.7 | 44 | 0 |
| Deaths | US_Ohio_Hamilton | 187 | 192 | 9.6 | 5 | 2.7 | 26 | 3 |
| Deaths | US_South_Dakota_Minnehaha | 54 | 55 | 2.1 | 1 | 1.9 | 29 | 0 |
| Deaths | Diamond_Princess | 13 | 12 | 0.0 | -1 | -7.7 | 22 | 0 |
| Cases | US_Texas_Harris | 23,047 | 21,967 | 9.8 | -1,080 | -4.7 | 12 | 727 |
| Deaths | India | 14,011 | 32,783 | 4.4 | 18,772 | 134.0 | 8 | 842 |
| Cases | Azerbaijan | 13,207 | 30,088 | 8.4 | 16,881 | 127.8 | 6 | 1324 |
| Cases | Panama | 26,752 | 38,757 | 8.5 | 12,005 | 44.9 | 7 | 1553 |
| Cases | US_California_Los_Angeles | 86,017 | 97,102 | 8.8 | 11,085 | 12.9 | 21 | 1629 |
| Cases | Russia | 591,465 | 703,979 | 7.9 | 112,514 | 19.0 | 36 | 2025 |
| Deaths | WholeWorld | 635,463 | 701,602 | 1.7 | 66,139 | 10.4 | 39 | 3482 |
| Cases | Egypt | 56,809 | 171,053 | 5.9 | 114,244 | 201.1 | 9 | 4947 |
| Cases | Saudi_Arabia | 161,005 | 337,073 | 5.7 | 176,068 | 109.4 | 6 | 5268 |
| Cases | Mexico | 185,122 | 754,786 | 2.6 | 569,664 | 307.7 | 14 | 5941 |
| Cases | Bangladesh | 115,786 | 267,358 | 5.8 | 151,572 | 130.9 | 8 | 9044 |
| Cases | Pakistan | 185,034 | 470,553 | 6.5 | 285,519 | 154.3 | 7 | 17969 |
| Cases | US | 2,312,302 | 2,089,628 | 9.8 | -222,674 | -9.6 | 35 | 21481 |
| Cases | India | 440,215 | 1,363,036 | 9.8 | 922,821 | 209.6 | 13 | 35566 |
| Cases | WholeWorld | 11,806,469 | 15,823,018 | 2.7 | 4,016,549 | 34.0 | 11 | 244799 |
| Cases | Iraq | 32,676 | 1 | 0.0 | -32,675 | -100.0 | - | - |
| Cases | Oman | 31,076 | 1 | 0.0 | -31,075 | -100.0 | - | - |
| Cases | Brazil | 1,106,470 | 1 | 0.0 | -1,106,469 | -100.0 | - | - |
| Cases | Argentina | 44,931 | 1 | 0.0 | -44,930 | -100.0 | - | - |
| Cases | Poland | 32,227 | 1 | 0.0 | -32,226 | -100.0 | - | - |
| Cases | Qatar | 88,403 | 1 | 0.0 | -88,402 | -100.0 | - | - |
| Cases | Chile | 246,963 | 1 | 0.0 | -246,962 | -100.0 | - | - |
| Cases | South_Africa | 101,590 | 1 | 0.0 | -101,589 | -100.0 | - | - |
| Cases | Bulgaria | 3,984 | 1 | 0.0 | -3,983 | -100.0 | - | - |
| Cases | Colombia | 71,367 | 1 | 0.0 | -71,366 | -100.0 | - | - |
| Cases | North_Macedonia | 5,196 | 1 | 0.0 | -5,195 | -100.0 | - | - |
| Cases | US_Florida_Broward | 11,327 | 1 | 0.0 | -11,326 | -100.0 | - | - |
| Cases | US_Nevada_Clark | 10,774 | 1 | 0.0 | -10,773 | -100.0 | - | - |
| Cases | Armenia | 20,588 | 1 | 0.0 | -20,587 | -100.0 | - | - |
| Cases | US_California_Alameda | 5,007 | 1 | 0.0 | -5,006 | -100.0 | - | - |
| Cases | US_California_Riverside | 13,800 | 1 | 0.0 | -13,799 | -100.0 | - | - |
| Cases | Senegal | 5,970 | 1 | 0.0 | -5,969 | -100.0 | - | - |
| Cases | US_Texas_Dallas | 17,299 | 1 | 0.0 | -17,298 | -100.0 | - | - |
| Cases | Bolivia | 25,493 | 1 | 0.0 | -25,492 | -100.0 | - | - |
| Cases | Kazakhstan | 18,231 | 1 | 0.0 | -18,230 | -100.0 | - | - |
| Cases | US_Florida_Palm_Beach | 10,943 | 1 | 0.0 | -10,942 | -100.0 | - | - |
| Cases | US_Georgia_DeKalb | 4,791 | 1 | 0.0 | -4,790 | -100.0 | - | - |
| Cases | US_North_Carolina_Mecklenburg | 8,956 | 1 | 0.0 | -8,955 | -100.0 | - | - |
| Cases | US_Arizona_Maricopa | 31,650 | 1 | 0.0 | -31,649 | -100.0 | - | - |
| Cases | US_Florida_Hillsborough | 5,973 | 1 | 0.0 | -5,972 | -100.0 | - | - |
| Cases | US_Texas_Bexar | 6,882 | 1 | 0.0 | -6,881 | -100.0 | - | - |
| Cases | US_Texas_Travis | 6,210 | 1 | 0.0 | -6,209 | -100.0 | - | - |
| Cases | Congo_Kinshasa | 5,924 | 1 | 0.0 | -5,923 | -100.0 | - | - |
| Cases | Honduras | 13,356 | 1 | 0.0 | -13,355 | -100.0 | - | - |
| Cases | US_Texas_Tarrant | 8,955 | 1 | 0.0 | -8,954 | -100.0 | - | - |
| Cases | Guatemala | 13,769 | 1 | 0.0 | -13,768 | -100.0 | - | - |
| Cases | Nigeria | 20,919 | 1 | 0.0 | -20,918 | -100.0 | - | - |
| Cases | US_Arizona_Pima | 5,587 | 1 | 0.0 | -5,586 | -100.0 | - | - |
| Cases | US_Tennessee_Shelby | 8,064 | 1 | 0.0 | -8,063 | -100.0 | - | - |
| Cases | Kenya | 4,797 | 1 | 0.0 | -4,796 | -100.0 | - | - |
| Cases | US_California_San_Bernardino | 9,361 | 1 | 0.0 | -9,360 | -100.0 | - | - |
| Cases | US_Ohio_Hamilton | 4,020 | 1 | 0.0 | -4,019 | -100.0 | - | - |
| Cases | US_California_Kern | 3,965 | 1 | 0.0 | -3,964 | -100.0 | - | - |
| Cases | US_Pennsylvania_Lancaster | 4,029 | 1 | 0.0 | -4,028 | -100.0 | - | - |
| Deaths | Iraq | 1,167 | 1 | 0.0 | -1,166 | -99.9 | - | - |
| Deaths | Egypt | 2,278 | 1 | 0.0 | -2,277 | -100.0 | - | - |
| Deaths | Pakistan | 3,695 | 1 | 0.0 | -3,694 | -100.0 | - | - |
| Deaths | Mexico | 22,584 | 1 | 0.0 | -22,583 | -100.0 | - | - |
| Deaths | US_Georgia_Fulton | 304 | 1 | 0.0 | -303 | -99.7 | - | - |
| Deaths | Colombia | 2,426 | 1 | 0.0 | -2,425 | -100.0 | - | - |
| Deaths | Chile | 4,502 | 1 | 0.0 | -4,501 | -100.0 | - | - |
| Deaths | North_Macedonia | 247 | 1 | 0.0 | -246 | -99.6 | - | - |
| Deaths | Saudi_Arabia | 1,307 | 1 | 0.0 | -1,306 | -99.9 | - | - |
| Deaths | Honduras | 395 | 1 | 0.0 | -394 | -99.7 | - | - |
| Deaths | US_California_Orange | 269 | 1 | 0.0 | -268 | -99.6 | - | - |
| Deaths | South_Africa | 1,991 | 1 | 0.0 | -1,990 | -99.9 | - | - |
| Deaths | Afghanistan | 598 | 1 | 0.0 | -597 | -99.8 | - | - |
| Deaths | Belarus | 351 | 1 | 0.0 | -350 | -99.7 | - | - |
| Deaths | Bolivia | 820 | 1 | 0.0 | -819 | -99.9 | - | - |
| Deaths | US_Washington_Yakima | 138 | 1 | 0.0 | -137 | -99.3 | - | - |
| Deaths | Azerbaijan | 161 | 1 | 0.0 | -160 | -99.4 | - | - |
| Deaths | Kazakhstan | 127 | 1 | 0.0 | -126 | -99.2 | - | - |
| Deaths | US_Minnesota_Ramsey | 211 | 1 | 0.0 | -210 | -99.5 | - | - |
| Deaths | Guatemala | 547 | 1 | 0.0 | -546 | -99.8 | - | - |
| Deaths | Oman | 137 | 1 | 0.0 | -136 | -99.3 | - | - |
| Deaths | Qatar | 99 | 1 | 0.0 | -98 | -99.0 | - | - |
| Deaths | Haiti | 88 | 1 | 0.0 | -87 | -98.9 | - | - |
| Deaths | Senegal | 86 | 1 | 0.0 | -85 | -98.8 | - | - |

At the moment of writing this manuscript, many countries or regions are still in the fast growing phase, and it is still impossible to predict the outcome of the epidemic in them (see **Table 3**). For others we can make predictions as shown in **Fig. 7**. Panels (a) & (b) show clearly that the BLP graph for Peru predicts a clear plateau for cases (*N*=478,000), but the predicted plateau for deaths is still rising rapidly. The plateau value for cases is almost double the current level of 257,000 making this a very meaningful forecast. Panel (c) shows that for Brazil the BLP predicts a stable plateau of 98,000, another very meaningful forecast, again almost double the current level of 47,000. Panel (d) shows that cases in Belarus are predicted to plateau at 82,000, although there is a less clear leveling. Panel (e) shows a split prediction for cases in the United Arab Emirates where there are two plateaus, at 49,000 and about 60,000, respectively; such splitting is very rare. Panel (f) shows that deaths in Kuwait are perhaps going to plateau at 400. Panels (a) to (c) are important forecasts with a meaningful impact, whereas those in panel (d) to (f) show the diversity of behavior making automatic forecasting a challenging problem.

**Figure. 7:**
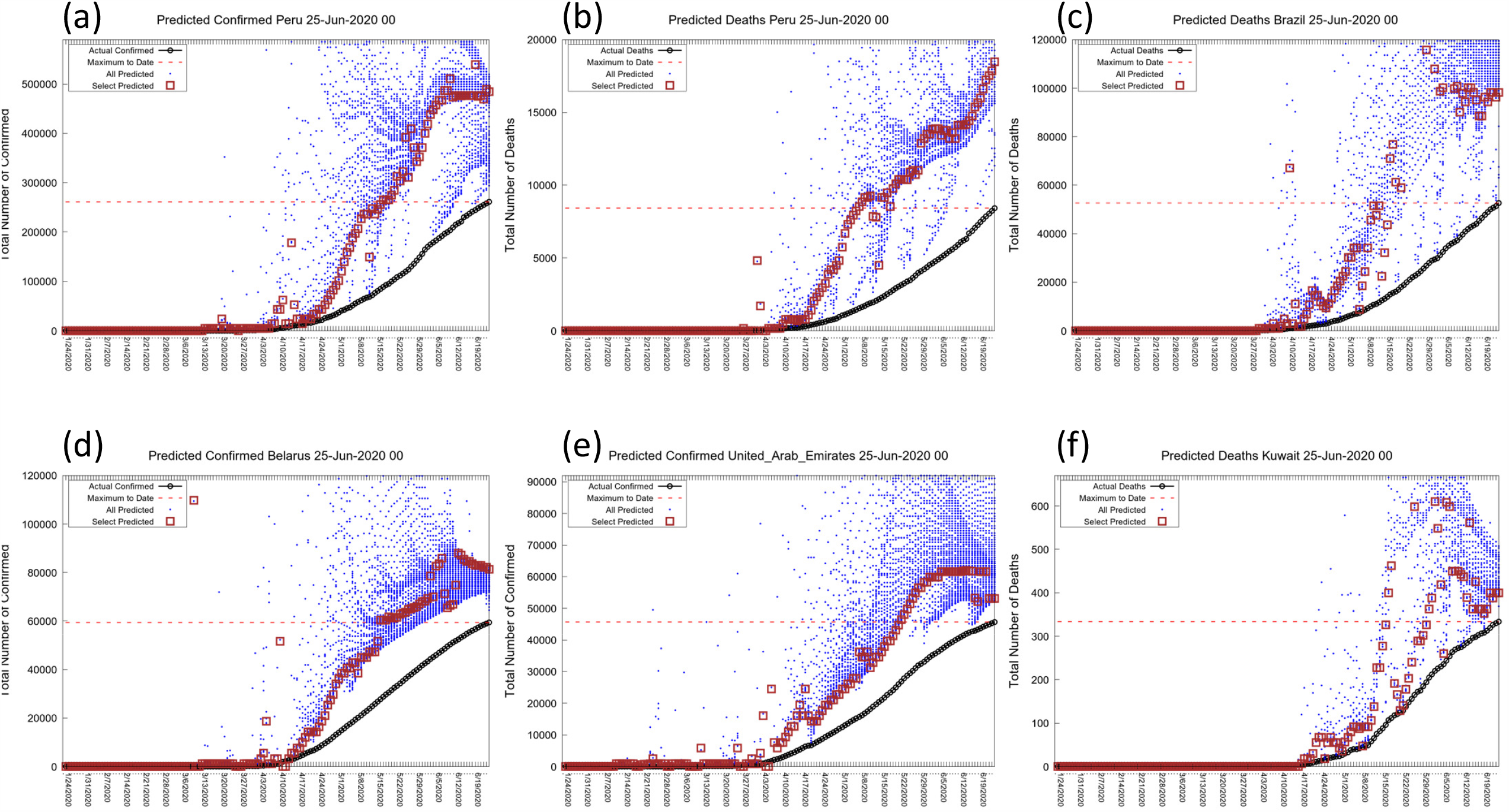
Best Line Predictions for Six Active Locations Ready for Forecast. Showing the BLP graphs for active locations with prediction approaching convergence of *N* (Peru, Brazil, Belarus, UAE, Kuwait)). These locations have been selected from **Table 3** because the predicted plateau is significantly higher than the current level (red dashed horizontal line). As this involves locations with large numbers of expected additional cases and deaths. Forecasting the outcome could be of major value to the countries involved. The locations also show a range of different behaviors.

### Open Availability of all Data

All graphs and tables are available online using apps written by Dr. Scaiewicz. The app at http://levitt1.herokuapp.com/ shows the classification for different countries and updated numbers and graphs. The app at http://levitt.herokuapp.com/ shows the predictions in the Best Line graphs. We thank Dr. João Rodrigues for advising us on this matter.

### Availability of the Computer Codes

We would like to make the computer codes we use available to all but these are currently written in a variety of languages that few would want to use. While Dr. Scaiewicz uses clean self-documenting Jupyter Python notebook code, Dr. Levitt still develops in a FORTRAN dialect call Mortran (Mortran 1975) that he has used since 1980. The Mortran preprocessor produces Fortran that is then converted to C-code using f2c. This code is at least a hundred-fold faster than Python code. His other favorite language is more modern, but involves the use of the now deprecated language Perl and Unix shell scripts.

Nevertheless, the methods proposed here are simple; they are easily and quickly implemented by a skilled programmer. Should there be interest, we would be happy to help others develop the code and test them against ours. We also realize that there is ample room for code optimization. Some of the things that we have considered are pre-calculating sums of terms to convert computation of the correlation coefficient from a sum over *N* terms to the difference of two sums. Another way to speed the code would be to use hierarchical step sizes in a binary search to find the value of ln*N* that gives the best straight line.

Our study involving as it did a small group working in different time zones and under extreme time pressure revealed that scientific computation nowadays faces a Babel of computer languages. In some ways this is good as we generally re-coded things rather than struggle with the favorite language of others. Still, we worry about the future of science when so many different tools are used. In this work we used Python for data wrangling and some plotting, Perl and Unix shell tools for data manipulation, Mortran (effectively C++) for the main calculations, xmgrace and gnuplot for other plotting, Excel (and Openoffice) for playing with data. And this diversity is for a group of three!

## DISCUSSION

### Non-Exponential Growth

It is evident from our data analysis that the growth of a COVID19 epidemic does not follow an exponential growth law even in the very first days, but instead its growth is slowing down exponentially with time. While all growth functions decelerate exponentially when approaching the plateau, the Gompertz function is unique in that it is decelerating from the first day, and thus can fit the first part of the COVID-19 outbreak. Moreover, its relatively simple functional form, allowed us to produce an efficient computer code to fit data in all different locations in a consistent way.

As would be expected, we find several examples in which this simple law is not followed, especially when looking at confirmed cases (deaths appear to follow the Gompertz Function more consistently). For some of these countries (e.g. Iran) it is evident that a second outbreak occurred well separated in time from the first. In other countries, (e.g. South Korea) we observe a change in the dynamics of the virus spread, which could be related to the adopted containment strategy or a difference in the level of testing. Even though such unusual dynamics cannot be predicted from the beginning, our fitting method is able to identify abrupt changes and will identify the slowest characteristic time and will, therefore, be able to produce a prediction for the new plateau.

We believe that the analysis in our study shows conclusively that COVID-19 epidemics grow according to the Gompertz Function and not the Sigmoid Function (Fig. 2). The main difference between these functional forms is that the Sigmoid Function starts off growing exponentially (it has a constant exponential growth factor) and then slows down (blue line in Fig. 2(c)). The Gompertz Function is never exponential but rather has a growth rate that decreases exponentially from the very first confirmed case. This does not make sense as when there are very few cases, it should be easy for each infectious individual to find people to infect, which would lead to exponential growth at the early stages of the outbreak. The Gompertz Function normally applies to conditions when the growth is constrained by some global resource. For example, bacteria growing with a limited food supply or a fire in an enclosure where oxygen is limited.

What is limited for coronavirus? First clues came from the large number of invisible cases indicated by the early serological studies by our Stanford colleagues (Bendavid 2020). More recently, a paper in Science (Silverman 2020) showed that millions of people were infected in the USA before there were known cases. The existence of invisible cases of individuals who are mildly symptomatic and, therefore, not counted as confirmed cases may explain the non-exponential behavior of COVID-19: the known cases cannot easily find people to infect as the hidden invisible cases have already infected them. We realize that other factors may limit growth. For example, the structure of the human interaction network can lead to sub-exponential growth (Moreno 2002). Still, we believe that as SARS-CoV-2 is so infectious, it does not have a problem finding people to infect early on due to the local network structure.

Initial sub-exponential growth is not a unique feature of COVID-19, but has been observed in previous viral outbreaks and needs to be taken into account to produce accurate predictions (Chowell 2016). Our method provides a quick way to analyze early epidemic data and identify and also quantify sub-exponential growth in terms of the time constant *U*.

### Clean and Curate Data Carefully

An essential step for our study has been to clean and curate the data made available from so many different countries. Had we not filled in missing value or spread large changes back in time, the sensitive methods we use would fail. Of course, we need to document every step we take so as not to manipulate data in some arbitrary way. In taking this approach we were aided by the fact that we started the project very early on when there were just 24 data points: six days of cases and deaths in two regions of China (Levitt 2020c).

Another consequence of being so intimately connected with the data is that we had to collect data manually until the various repositories became established. We are now quite certain that the quality of data is more or less the same from all sources. The question of data reliability is often raised and we believe that the data has to obey so many rules of self-consistency that cheating would be almost impossible. For example, in Fig. 2, we see that the raw data from China, non- Hubei. which was available in late January is essentially indistinguishable from the data released for New Zealand two months later.

### Sanity Tests To Prove We Do Not Inadvertently Cheat

In a study like this involving a huge body of data, computer programs written quickly and the intense pressure to get results out while they can still be useful, one needs to be very self-critical at every stage looking for computer bugs that could explain any good results that one finds. Specifically, we are trying to test our forecasting method by going back in time and trying to predict something that was not known then but is know now. Such a process, often called ‘postdiction’ in contrast to ‘prediction’, is extremely dangerous. We guard against it by running calculations with data sets that have been specially prepared to eliminate all data after a certain previous date. This is tricky in that one cannot use smoothed data as smoothing looks into the future to smooth the present. In this work we made a series of data sets going back into the past and showed that the results from a past date would have been obtained with a data set that did not include data after that date.

### Work in progress

We have been studying COVID for five months and worked on all aspects of the analysis.

Some of the related projects that we are working on include:

a. Predict the future time-course of the epidemic and not just the plateau value *N*. This will involve better understanding of the two other parameters of the best line fit, *U* & *T*.
b. In what ways are the detailed trajectories from various locations different? What affects the trajectory in terms of *N* and *U*: population size, population age/health, physical size of location, social distancing or lockdown measures?
c. What is the burnout saturation value of *N*? What is the population fatality ratio if the infection runs its course?

## CONCLUSIONS

This manuscript is being submitted as a preprint, which is something that we have never done before. We do this for two reasons. One is to make our discoveries available to all at a stage where they will still be useful. Another is to solicit broad criticism and comments that are essential to the scientific process.

## Data Availability

All data is to be made available

http://levitt.herokuapp.com/

## ACKNOWLEDGMENTS

This analysis started as a team effort based on our 14-Mar-20 analysis (Levitt 2020b); besides the current authors, it involved Patrick Tam (Hong Kong, see www.covibes.org), Frédéric Poitevin, João Rodrigues and Fatima Pardo-Avila (all at Stanford). We also had discussions and shared ideas with Eran Bendavid (Stanford), Cathrine Bergh (Royal Institute of Technology, Sweden) and Siri Camee van Keulen (Utrecht University, The Netherlands). We are most grateful to them. We offer special thanks to João Rodrigues, for his automatic daily data updates and invaluable advice on writing Python apps.

This work was supported by a US National Institutes of Health award R35GM122543 to M.L. and by a National Natural Science Foundation of China Grant No. 31770776 to F.Z. Michael Levitt is the Robert W. and Vivian K. Cahill Professor of Cancer Research.

## ONLINE RESOURCES USED OR CITED IN THE TEXT

(Rajkumar) https://www.kaggle.com/allen-institute-for-ai/CORD-19-research-challenge

(JHCS) https://gisanddata.maps.arcgis.com/apps/opsdashboard/index.html#/bda7594740fd40299423467b48e9ecf6

(JOBTUBE) https://jobtube.cn/wv/?from=groupmessage&isappinstalled=0

(JHU) https://s3-us-west-1.amazonaws.com/starschema.covid/JHU_COVID-19.csv

(Starschema) https://starschema.com/covid-19-data-set

(Ita-regioni)https://raw.githubusercontent.com/pcm-dpc/COVID-19/master/dati-regioni/dpc-covid19-ita-regioni.csv

(Levitt-Twitter) https://twitter.com/MLevitt_NP2013/status/1256511516863586304?s=20

## LEGENDS FOR SUPPLEMENTARY FIGURES

**Figure S1.**
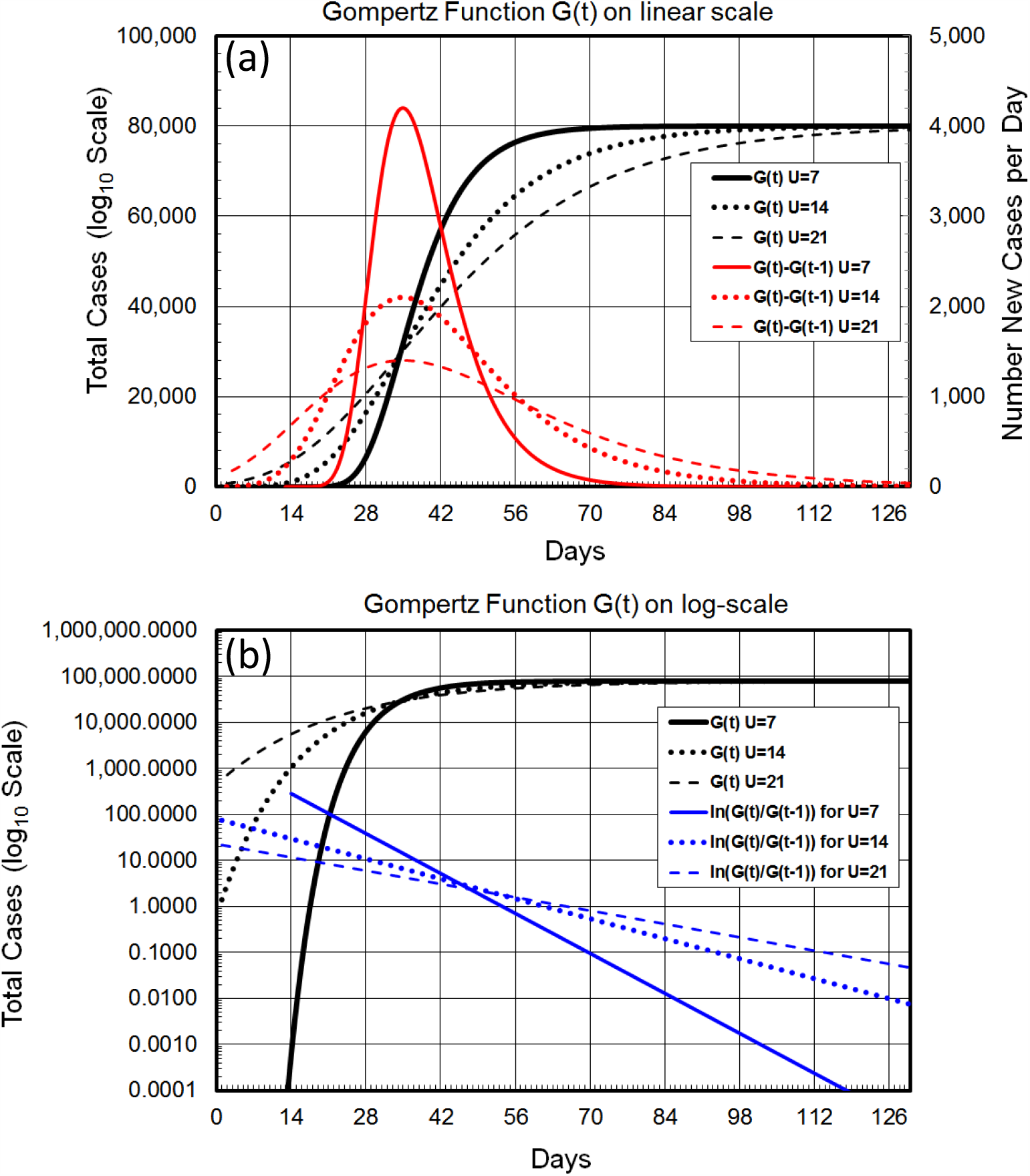
Showing affect of the parameter U (in days) on the Gompertz Function. Showing how the *U* parameter has a major effect on the shape of the Gompertz Function, affecting as it does the trajectory of the Total Count (*X(t)*, in black), the new counts by day ((*X(t)-*(*X(t-1)*, in red) and the gradient of the ln(Total Counts), which is ln[*X(t)*]-ln[*X(t-1)*] or ln[ln[*X(t)/X(t-1)*]] (in blue). The solid lines show trajectories for the Total Counts, New Counts, and Gradient (*H(t)*) for a *U* parameter of 7 days, the shortest decay time seen for real cases (Table 2). The dotted lines show the same data for *U*= 4 days and the dashed line shows the same data for *U*=21 days. The trajectory of ln[*H(t)*], the gradient of ln[*X(t)*], is a simple straight line with slope of *1/U*

**Figure S2.**
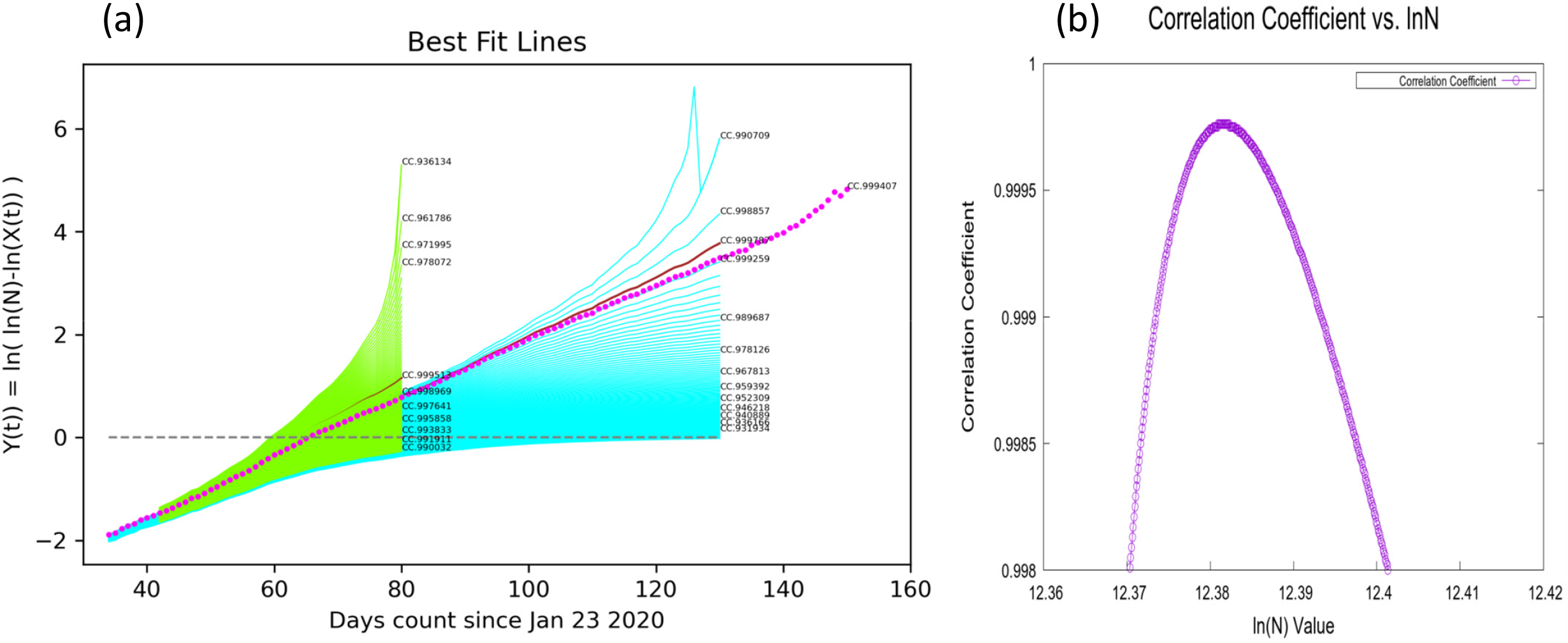
Showing how the line *Y(t)=ln[ln[N/X(t)]]* varies for different values of *N*. The straightness of the line is measured by the correlation coefficient between *Y(t)* and *t*. The value of the correlation coefficient varies smoothly as value of *N* is changed. (a) Showing how straight line fits have strong predictive power. The lines in green are fitted to data that was available 50 days ago. The line in magenta is fitted to current data and is a straight-line continuation of the best line 50 days ago. The Correlation Coefficient (*CC*), which is used to measure the straightness of the line as a function of ln(N), is sensitive to departure from linearity. (a) Shows that as ln(*N*) varies the *CC* value reaches its maximum smoothly. (b) Distribution of correlation coefficients value as the guessed value of ln(*N*) is changed.

**Fig S3.**
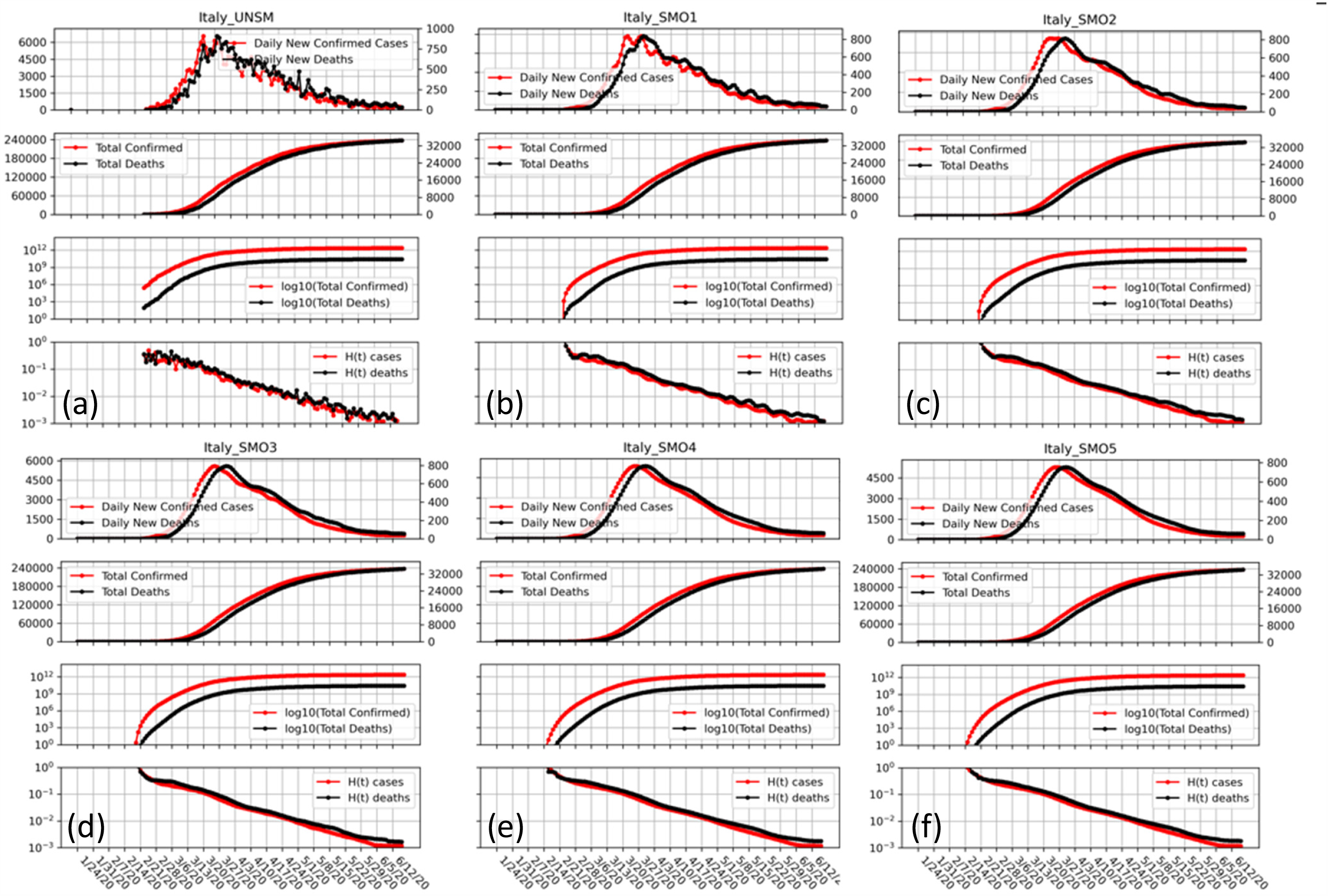
Showing five levels of smoothing on our standard four panel plots. Showing our Four-Panel graphs with different levels of smoothing of the data using the LOWESS method (see text). The strength of the smoothing increasing progressively for SMO1 through SMO5 and one sees that while local ripples are eliminated there is no shift of the peak position. Such shift do occur with simpler smoothing schemes such a running averages.

## Supplementary Methods

Proof that any growth function has a *Y(t)* function that tend to be linear for large *t*.

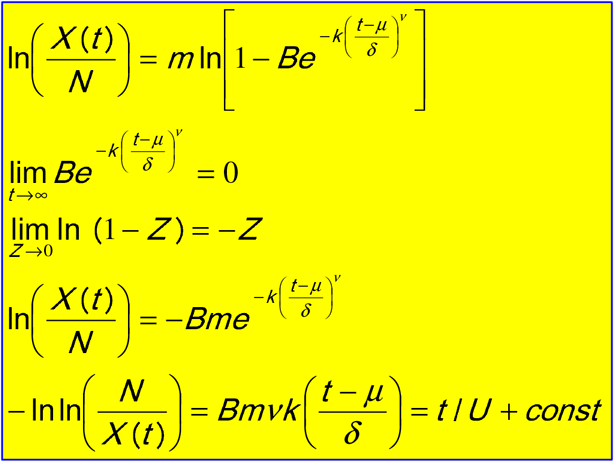

